# *TLR8* escapes X chromosome inactivation in human monocytes and CD4^+^ T cells

**DOI:** 10.1101/2023.08.08.23293823

**Authors:** Ali Youness, Claire Cenac, Berenice Faz-López, Solange Grunenwald, Franck J. Barrat, Julie Chaumeil, José Enrique Mejía, Jean-Charles Guéry

**Affiliations:** Institut toulousain des maladies infectieuses et inflammatoires (INFINITY), Université de Toulouse, UMR 1291 INSERM, CNRS, Toulouse, France; Service d’Endocrinologie, Maladies Métaboliques et Nutrition, Hôpital Larrey, Centre Hospitalier Universitaire (CHU) de Toulouse, Toulouse; HSS Research Institute and David Z. Rosensweig Genomics Research Center, Hospital for Special Surgery, New York, NY; Department of Microbiology and Immunology, Weill Cornell Medical College of Cornell University, New York, NY; Université Paris Cité, Institut Cochin, INSERM, CNRS, F-75014 PARIS, France

## Abstract

Human endosomal Toll-like receptors TLR7 and TLR8 recognize self and non-self RNA ligands, and are important mediators of innate immunity and autoimmune pathogenesis. TLR7 and TLR8 are encoded by the adjacent X-linked genes, *TLR7* and *TLR8*. We previously established that *TLR7* evades X chromosome inactivation in female immune cells, and that mononuclear blood cells express more TLR7 protein in women than in men. Using RNA fluorescence *in situ* hybridization, we now show that *TLR8* likewise evades X chromosome inactivation in CD14^+^ monocytes and CD4^+^ T lymphocytes, and that cells harboring *TLR7* or *TLR8* transcript foci are more frequent in women than in men. In parallel, we found *TLR7* and *TLR8* simultaneous transcription to be disproportionally frequent in female monocytes and T cells, and disproportionally scarce in the male cells, resulting in a 7-fold difference in frequency. These transcriptional biases were again observable when comparing the single X of XY males with the active X of female cells. Among (47,XXY) Klinefelter syndrome males, both *TLR7* and *TLR8* escape X chromosome inactivation, and co-transcription frequencies on the active X of monocytes were intermediate overall between those for XY males and XX females, and encompassed both male- and female-like individual patterns. These findings indicate that the *TLR7* and *TLR8* genes form a co-regulated gene cluster, which we have called the X-linked Toll-like receptor locus, with different sex- and sexual karyotype-dependent modes of transcription. Interestingly, TLR8 protein expression was significantly higher in female mononuclear blood cells, including all monocyte subsets, than in the male cells. Thus, co-dependent transcription from the active X chromosome and escape from inactivation could both contribute to higher TLR8 protein abundance in female cells, which may have implications for the response to viruses and bacteria, and the risk of developing inflammatory and autoimmune diseases.

**Highlights:**

- *TLR8*, like *TLR7*, escapes X chromosome inactivation in immune cells from women and 47,XXY Klinefelter syndrome (KS) men.
- The frequency of cells double-positive for *TLR7* and *TLR8* primary transcripts is 7-fold higher in women than in men.
- *TLR7* and *TLR8* form a co-regulated gene cluster on the human X chromosome, with sex-specific, divergent transcriptional patterns observable in monocytes and CD4^+^ T lymphocytes.
- Co-dependent transcription of the *TLR7* and *TLR8* genes on the active X was observed in women and KS men, contrasting with mutually exclusive transcription in euploid men.
- Blood mononuclear cells, including monocyte subsets, expressed higher levels of TLR8 protein in females than in males.

## Introduction

Human Toll-like receptors 7 and 8 (TLR7, TLR8) are essential components of the innate immune response to microbial pathogens. These paralogue receptors recognize RNA degradation products from viruses, intracellular bacteria, fungal and protozoan pathogens, and also endogenous sources (1–11). TLR7 and TLR8, along with receptors for CpG-unmethylated DNA (TLR9) and double-stranded RNA (TLR3), make up the subfamily of endosomal nucleic acid-binding TLRs (12). Crystal structures have revealed TLR7 and TLR8 to be dual sensors possessing two ligand-binding sites working in synergy, one for small agonists, namely guanosine in TLR7 and uridine in TLR8, and a separate site for short single-stranded RNA molecules: uridine-rich RNA in TLR7, and U- and G-containing RNA in TLR8 (13–15).

TLR7 and TLR8 exhibit different expression landscapes among human leukocytes (16, 17). TLR7 is found primarily in plasmacytoid dendritic cells (pDCs), monocytes, and B lymphocytes, whereas TLR8 is preferentially expressed in monocytes, myeloid dendritic cells, and neutrophils. Vigorous production of type I interferon (IFN) by pDCs upon TLR7 stimulation is a key component of the antiviral response, whereas TLR7 and TLR8 signaling leads to the secretion of proinflammatory cytokines in cells of the monocyte/macrophage lineage. TLR7 engagement, however, is also known to elicit anergy and cell death in CD4^+^ T cells during chronic infection (18, 19), and TLR8 engagement is reported to reverse Foxp3^+^ Treg cell function through inhibition of glucose uptake and glycolysis (20, 21). Accordingly, ligand sensing by TLR7 or TLR8 initiates a variety of processes across immune cell populations, as illustrated by single-stranded viral RNA fragments from SARS-CoV-2, which elicit either an antiviral or a proinflammatory response in pDCs, myeloid dendritic cells and lymphocytes (22, 23).

Human endosomal TLRs have evolved under stringent purifying selection owing to their non-redundant role in preserving host fitness (24), and only of late have null mutations of the X-linked genes, *TLR7* and *TLR8* been discovered. During the spread of SARS-CoV-2, deficiency in type I interferon-dependent antiviral immunity caused by rare loss-of-function or hypomorphic *TLR7* mutations emerged as a determinant of life-threatening COVID-19 in men under 60 (25–29), even though TLR7 engagement in pDCs may also promote the macrophage-induced cytokine storm in COVID-19 patients (23). Excess TLR7 or TLR8 activity, by contrast, can lead to sterile inflammation and autoimmunity, and is an important contributor to the pathogenesis of autoimmune syndromes (30, 31). Its role in the development of autoimmunity was established with the aid of murine models, where the expression of two copies of *Tlr7* or *Tlr8* was sufficient to induce lupus-like manifestations (16, 31–34). Introgression of TLR7 deficiency into genetic backgrounds predisposing to lupus conversely protected the mice against autoimmunity (35, 36). Remarkably, dendritic cells of TLR8-deficient mice overexpressed *Tlr7* and were hyperresponsive to TLR7 ligands, and the animals developed autoimmunity (37, 38). These animal models prefigured the phenotypes for human mutations that have been described only recently. A missense mutation (p.Tyr264His) increasing TLR7 affinity for guanosine was carried by a female child suffering from severe lupus, and this dominant gain-of-function variant of *TLR7* was sufficient to cause autoimmunity in both male and female transgenic mice (39). Another study identified a missense *TLR8* mutation leading to partial TLR8 deficiency in monozygotic twin boys with severe autoimmune hemolytic anemia and a TLR7-dependent autoinflammatory phenotype (40). These human and animal studies call attention to *TLR7* and *TLR8* as bidirectionally dosage-sensitive genes underpinning a tonic level of immune activity between the extremes of defective antiviral defense and severe autoimmune pathogenesis.

The *TLR7* and *TLR8* genes lie within a narrow, 56-kb interval on the short arm of chromosome X (**Fig 1A,B**). These paralogue genes arose from an ancestral autosomal gene that duplicated in the vertebrate line before the divergence of tetrapods from fishes ∼400 million years ago (41, 42). In eutherian mammals, one of the two X chromosomes of female cells is randomly inactivated during early embryonic development to equalize the dosage of gene expression between the sexes. An early step of this process is the coating in *cis* of the X chromosome to undergo inactivation by the long noncoding RNA, XIST (43). In men with Klinefelter syndrome, who carry an aneuploid karyotype encompassing one or more supernumerary X chromosomes, all but one X are similarly inactivated (44). Around 15% to 23% of human X-linked genes escape X chromosome inactivation (XCI) to a variable extent. Some of these genes escape XCI in a constitutive manner, whereas a majority are defined as facultative escapees as they evade XCI in certain cells, in some tissues or in some individuals (45–47). We previously demonstrated that the *TLR7* gene can escape XCI and is therefore transcribed in bi-allelic fashion in a fraction of female immune cells in all individuals tested (48, 49) but also in immune cells from 47,XXY Klinefelter syndrome (KS) males (48). Interestingly, KS men have an equivalent risk to women to develop relatively rare immune disorders such as systemic lupus erythematosus (SLE) (50), Sjogren’s syndrome (51) or Systemic Sclerosis (SSc) (52), suggesting a dominant role of X-linked genetic effects over sex hormones in autoimmune disease susceptibility. Whether *TLR8* also evades XCI, however, has not yet been explored. The close proximity between the two genes, and the homology and similarity of function between the encoded receptors, warranted investigating whether *TLR8*, mirroring *TLR7*, could be transcribed on the inactive X chromosome (Xi) of the immune cells of women and Klinefelter syndrome men.

**Figure 1.**
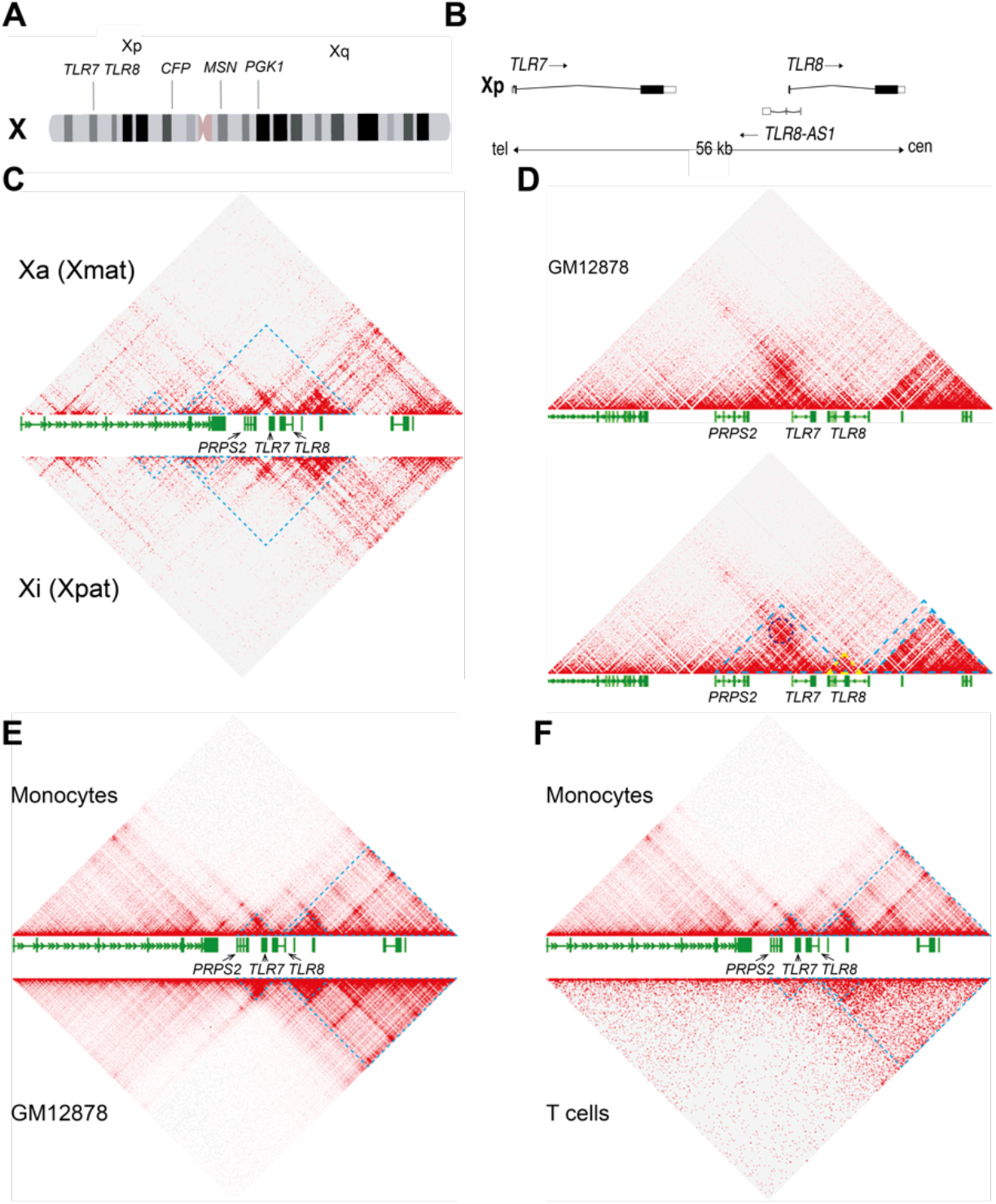
3D conformation of the *TLR7-TLR8* genomic region in male and female cells. (**A**) Idiogram of the human X chromosome with the positions of the adjacent genes, *TLR7* and *TLR8*, and three marker genes, *CFP*, *MSN and PGK1*, that do not evade X chromosome inactivation. (**B**) Map of the *TLR7*, *TLR8-AS1* and *TLR8* locus on Xp22.2; tel, cen denote the telomeric and centromeric ends of the map.(**C**) Hi-C map of interactions around the *TLR7-TLR8* region on the active (Xa, top panel) or inactive (Xi, bottom panel) X chromosome at 5 kb-resolution in GM12878 human female cells. (**D**) Hi-C map of interactions around the *TLR7-TLR8* region at 1 kb-resolution in GM12878 human female cells (datasets in **C** and **D** from Rao et al, 2014). The dark blue circle denotes a peak of interaction (loop) between *TLR7* and *PRPS2* as determined by the Peak tool in the Juicebox software. Blue and yellow dashed lines represent domains of preferential interactions as defined by the “Contact Domain” tool in the Juicebox software. (**E, F**) Hi-C maps of interactions around the *TLR7-TLR8* region at 5-kb resolution in human male monocytes (**E, F**), top panels; dataset from Phanstiel et al, 2017) versus GM12878 human female cells (**E**, bottom panel, dataset from Rao et al, 2014) or male T cells (**F**, bottom panel, dataset from Zhang et al, 2020). Blue dashed lines represent domains of preferential interactions as defined by the “Contact Domain” tool in the Juicebox software.

We have studied here CD14^+^ monocytes and CD4^+^ T lymphocytes, where both TLR7 and TLR8 are expressed (18, 53–55), using RNA fluorescence *in situ* hybridization (RNA-FISH) to visualize on a single-cell basis the primary transcripts of *TLR7* and *TLR8* relative to X chromosome territories. We provide evidence that *TLR8,* like *TLR7,* escapes XCI in CD4^+^ T cells and monocytes. In addition, we have gathered evidence for co-dependent transcription of the *TLR7* and *TLR8* genes on the active X chromosome (Xa) of the cells from women and KS men, in a comparison with euploid men (46,XY). Importantly, TLR8 protein expression was found to be up-regulated in female immune cells, including monocytes, suggesting that both XCI escape and co-dependent transcription could enhance TLR8 protein abundance in females.

## Materials and methods

### Donors and ethical compliance

This study complied with the ethical principles of the Declaration of Helsinki, and with applicable French regulations. Peripheral blood mononuclear cells (PBMCs) of anonymous, healthy blood donors at the Toulouse blood transfusion center (Etablissement Français du Sang) were from an in-house biobank approved by the competent ethics board (Comité de Protection des Personnes Sud-Ouest et Outre-Mer II, Toulouse) under reference 2-15-36. Klinefelter syndrome males presented with a 47,XXY karyotype, and were aged 16 to 44 at the time of sample collection. These patients were enrolled at the Toulouse University Hospital, and their study was approved by the aforementioned ethics board under reference 1-16-28. Written consent was obtained from each patient or, for child participants, from the legal guardian.

### Isolation of CD14^+^ monocytes and CD4^+^ T lymphocytes

Upon thawing, PBMCs from healthy (female n=6; males n= 7) or Klinefelter syndrome donors (n=5) were cultured overnight in RPMI 1640 medium supplemented with 2 mM L-glutamine, 0.5 mM sodium pyruvate, 5 mM HEPES, and non-essential amino acids (all from Life Technologies); 100 U/ml penicillin and 100 µg/ml streptomycin (BioWhittaker); 5% human AB serum (Gemini Bio-Products); and, for T-cell work, 10 ng/ml human recombinant IL-2. Classical CD14^+^ monocytes and CD4^+^ T cells were respectively isolated with EasySep Human CD14 Positive Selection and Human CD4 Negative Selection kits from STEMCELL Technologies. Monocytes were cultured in 96-well polypropylene plates to avoid non-specific activation arising from adhesion to polystyrene. Naive CD4^+^ T cells were seeded into 24-well plates (10^6^ cells/well), and stimulated for three days in the presence of 10 ng/ml recombinant human IL-2 and 12.5 µl/ml ImmunoCult Human CD3/CD28 T Cell Activator (STEMCELL Technologies).

### Flow cytometry

Human immune cells were stained with fluorescent mouse monoclonal antibody (mAb) conjugates against specific surface markers to study CD4^+^ T cell activation: PE-Cy5 anti-human CD69, clone NF50; and BV421 anti-human CD25, clone M-A251 (both from from BD Biosciences). Flow cytometry analysis was performed on a BD Biosciences LSR II flow cytometer or FACSAria II cell sorter, and the data were processed with the FlowJo software (FlowJo LLC).

For intracellular TLR8 staining, freshly thawed PBMCs (5 × 10^6^ cells/ml) from age-matched male and female healthy subjects from our in-house biobank were surfaced stained with PE-Vio615 conjugated Lin-specific mAb (anti-CD3, cloneBW264/56; anti-CD19, clone LT19; and anti-CD56, clone REA196; all from Miltenyi Biotec), anti-CD14-PB (clone REA599, Miltenyi Biotec) and anti-CD16-AF700 (clone 5C3, BD Biosciences). Cells were then fixed and intracellularly stained using fixation and permeabilization buffers (#00-5123-43 and #00-8333-56, from eBioscience) with anti-TLR8-PE and anti-TLR8-APC mAb (clone S16018A, BioLegend). Flow cytometry analysis was performed on a BD Biosciences Fortessa instrument, and monocyte subsets defined as the Lin^−^ CD14^+^CD16^−^ (classical monocyte), CD14^+^CD16^+^ (intermediate monocyte), and CD14^−^CD16^+^ (non-classical monocyte). Data were processed with the FlowJo 10.9 software.

### Quantification of TLR8 protein expression by Western blotting

PBMCs were thawed and cultured at 37°C for 2 hours before counting, cell lysate preparation, and western blotting as below. Cell lysates were prepared in Laemmli sample buffer (Invitrogen), sonicated, and total protein quantified by a bicinchoninic acid protein assay (Pierce). Samples were heated for 10 minutes at 70°C in the presence of a reducing agent (Invitrogen). Protein (20–25 µg per lane) were fractionated by SDS-PAGE on precast 4%–15% gradient Stainfree gels (Bio-Rad), and transferred to Amersham Hybond 0.45-µm PVDF membranes (GE Healthcare). The membranes were blocked with 5% skim milk, 0.1% Tween-20 in PBS, probed overnight with the anti-N-terminus (LRR1)-specific anti-human TLR8 mAb (rabbit monoclonal IgG, clone D3Z6J #11886, Cell Signaling Technology), and finally incubated with suitable peroxidase-conjugated secondary antibodies (anti-rabbit IgG, #7074, Cell Signaling Technology). Chemiluminescent detection was carried out with Amersham ECL Prime reagent (GE Healthcare), and densitometric analysis performed with the Image Lab 5.0 software (Bio-Rad). TLR8 protein quantification was performed as described (56). The densitometric signals of the full-length TLR8 120-kDa forms were normalized to total protein and then to an internal standard PBMC lysate that was loaded in each gel for inter-gel data normalization as described (56).

### RNA FISH Probes

Probes for RNA FISH were prepared by PCR amplification of human genomic DNA fragments using the primer sets in **S1 Table**, targeting exon 1 of *XIST* (MIM *314670), and both exon and intron regions of *TLR7* (Xp22.2; MIM *300365), *TLR8* (Xp22.2; MIM *300366), *CFP* (Xp11.23; MIM *300383), *MSN* (Xq12; MIM *309845) and *PGK1* (Xq21.1; MIM *311800). The *XIST* and *TLR7* probes have been described previously (48). To exclude repetitive DNA from the PCR amplimers, genomic sequences were filtered *in silico* with the RepeatMasker tool (57) prior to primer design. Probes were fluorescently labeled using the Vysis Nick Translation kit (Abbott) according to the manufacturer’s instructions, and any of the following dUTP conjugates: aminoallyl-dUTP-ATTO-655, aminoallyl-dUTP-ATTO-550, or aminoallyl-dUTP-XX-ATTO-488 (all from Jena Bioscience). The Xa marker genes, *CFP*, *MSN* and *PGK1*, were identified as non-escape genes in the scientific literature (47, 58), supported by additional molecular genetics information from the OMIM database at https://omim.org/, and monocyte expression data from the EBI-EMBL Expression Atlas at https://www.ebi.ac.uk/gxa/home.

### RNA FISH

RNA FISH was performed as described in our earlier reports (48, 59). Briefly, spreads of monocytes or T lymphocytes on poly-L-lysine-coated coverslips were fixed for 10 min with 3% paraformaldehyde at room temperature, and permeabilized for 7 min in ice-cold cytoskeletal buffer containing 0.5% Triton X-100 and 2 mM vanadyl-ribonucleoside complex (New England Biolabs). The cells were dehydrated through successive ethanol baths, air-dried briefly, and incubated with the labeled probes overnight at 42°C. The coverslips were rinsed twice with 50% formamide in 2× SSC (saline sodium citrate) and thrice with 2× SSC alone, and nuclei were counterstained with DAPI in phosphate buffered saline. The coverslips were slide-mounted using Dako fluorescence mounting medium before microscopy on a Leica TCS SP8 or Zeiss LSM710 microscope using a 63× oil immersion objective. Image data were processed with the Fiji software (https://fiji.sc/).

### Cell scoring

In RNA FISH experiments, individual cells were scored as positive or negative for the gene of interest depending on the detection or non-detection of primary transcript foci under microscopic examination. To quantitate the escape from XCI for *TLR7* or *TLR8*, we counted the cells displaying either bi-allelic transcriptional foci or a single signal on the Xi, and expressed the percentage of escape cells with reference to the number total of cells positive for the gene under consideration. In estimating the overall frequencies of *TLR7*, *TLR8*, and joint *TLR7* and *TLR8* transcriptional foci, cells were scored by two alternative approaches: either (i) in random microscopic fields, by considering all cells regardless of signals from the Xa marker probe, or (ii) by including only those cells marked by the Xa probe.

### Statistical analysis

For *TLR7* and *TLR8* considered together, cell counts were cross-classified on a per-donor basis as a 2×2 contingency table with *TLR7* and *TLR8* signals as nominal variables with positive or negative outcomes as above. Statistical analysis was then performed in the R computing environment (60). From each 2×2 table classifying *N* cells total,

|  |  |  |
| --- | --- | --- |
|  | *TLR7*<sup>+</sup> | *TLR7*<sup>-</sup> |
| *TLR8*<sup>+</sup> | *a* | *b* |
| *TLR8*<sup>-</sup> | *c* | *d* |

we derived descriptive statistics, i.e., the percentage of scored cells in a stratum of interest, and also a measure of association between *TLR7* and *TLR8* transcriptional signals, namely Yule’s Q coefficient of association in 2×2 tables (61) as given by equation 1.

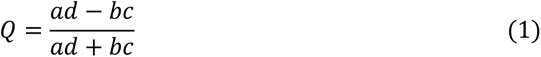

In parallel, we considered the ratio of the observed frequency of *TLR7*^+^ *TLR8*^+^ cells to the frequency expected under the null hypothesis of independent transcription of the two genes. We computed this *obs*/*exp* ratio according to equations 2 and 3, where the probability *p*_7·8_ of observing transcriptional foci for *TLR7* and *TLR8* together is given, under independence, by the product of the respective probabilities of observing *TLR7* and *TLR8* signals.

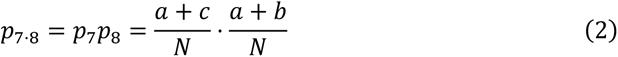

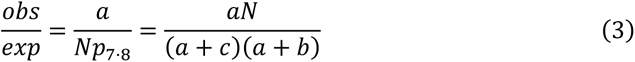

We summarized the RNA FISH data for a given group by a statistical procedure built around R library *gmeta* (62), which performs meta-analysis based on the mathematics of confidence distributions (CDs) (63). We generated a CD for each statistic under consideration and for each individual, and fed the individual CDs together to the *gmeta* function for summarization under a random effects model. CDs for a proportion (such as percent cells evading XCI, or positive for both *TLR7* and *TLR8* transcripts) were computed by a binomial approach; CDs for the analytical statistics, Yule’s Q and the *obs*/*exp* ratio, were estimated non-parametrically by bootstrap resampling with 10000 or 50000 replications. The software computed a point estimate and 95% CI from each individual CD, and pooled the individual CDs into a summary CD to derive a group’s meta-analytical mean and its 95% CI.

To test for independence in 3×2 tables encompassing sparse cells (**S5 Data**), we performed χ^2^ tests in R by a Monte Carlo procedure with 10^6^ replications. When carrying out between-groups comparisons, e.g., males versus females, we computed a p-value to test the difference between the group summaries for the statistic under consideration. For this, we determined the probability for the group-A meta-analytical mean according to the CD for group B, and the probability for the group-B meta-analytical mean according to the CD for group A; we took the greater of the two one-tailed p-values, and multiplied it ×2 to approximate a two-tailed hypothesis test. We computed a similar, two-tailed CD-based p-value to test whether a group’s summary for a measure of association (Yule’s Q, or the *obs/exp* ratio) diverged from the null value denoting independence, by performing a single comparison between the corresponding null value (Q = 0, or *obs/exp* = 1) and the group’s summary CD for this statistic. Our R scripts performing two- and three-group meta-analytical summarization, hypothesis tests, and plotting are available from the Zenodo repository under digital object identifiers (DOIs) https://doi.org/10.5281/zenodo.6580369 and https://doi.org/10.5281/zenodo.6580378. We performed the Pearson correlation analyses in **Fig S4** with GraphPad Prism 7.0a from GraphPad Software.

### Hi-C dataset analysis

We used the Juicebox software (64), including the Contact Domains and Peaks calling tools, at http://www.aidenlab.org/juicebox/ to access and visualize the following chromatin conformation datasets: (i) data for human female lymphoblastoid cell line GM12878 from Rao and coll. (65), Gene Expression Omnibus (GEO) accession GSE63525; (ii) data for male human monocytic cell line THP-1 from Phanstiel and coll. (66), Sequence Read Archive (SRA) accession PRJNA385337; and (iii) data for non-activated, human primary CD3^+^ T cells from Zhang and coll. (67), GEO accession GSE104579.

## Results

### 3D conformation of the *TLR7-TLR8* region on the X chromosome

We first investigated chromatin conformation over the *TLR7-TLR8* region of chromosome X by leveraging available chromosome conformation capture (Hi-C) datasets (64, 68). Data from the GM12878 human transformed lymphoblastoid female cell line allowed allelic discrimination of the conformations of the Xa, of maternal origin in this line, and the paternal Xi (65). The overall conformation of X chromosomes confirmed the erosion of topologically associating domain (TAD) structures on the Xi, substituted as expected by two mega-domains (data not shown), but no major conformational differences were observed at 5kb-resolution between the Xa and the Xi over the *TLR7-TLR8* region, confirming that escape regions retain the typical TAD topology (69) (**Fig 1C and Fig S1).** This suggested that global non-allelic Hi-C analyses of female X chromosomes can be informative about the typical conformation of the *TLR7-TLR8* region in both the Xa and Xi, enabling a comparison with the single X chromosome of male cells.

High 1kb-resolution, non-allelic Hi-C data from GM12878 cells (65) revealed that *TLR7* and *TRL8* are located in the same domain of interactions (**Fig 1D**). However, on a close look at this region, it is striking that the *TLR7* promoter makes a strong interaction with the adjacent *PRPS2* gene, denoted by a characteristic loop signal (**Fig 1D**, dark blue circle), whereas *TLR8* seems to form a sub-domain of its own (**Fig 1D**, yellow triangle). Similar conformations were found in male monocytes (66) and T cells (67) (**Fig 1E,F**). These data on the 3D conformation of the *TLR7-TLR8* genomic region suggested different patterns of transcriptional regulation for *TLR7*, on one hand, and for *TLR8*, on the other, and prompted us to examine the expression of primary transcripts from either gene using RNA FISH on immune cell types where both receptors were known to be co-expressed.

### *TLR7* and *TLR8* evade X chromosome inactivation in female monocytes and CD4+ T cells

We used RNA FISH probes to detect both *TLR7* and *TLR8* primary transcripts on the Xa and Xi of CD14^+^ monocytes and CD4^+^ T lymphocytes from women. To discriminate the Xa and Xi chromosomal origins of *TLR7* or *TLR8* primary transcripts, early RNA FISH experiments involved a previously validated probe for the long non-coding RNA XIST (reference (48), and **Table S1**), but this probe failed to paint the Xi territory in female monocytes (not shown). Similar occurrences of XIST non-detection have been noted by others with regard to resting B and T lymphocytes (70–72). Our alternative strategy was to differentiate instead the Xa of female cells. For this, we searched the scientific literature and relevant databases for X-linked genes subject to XCI that were well-expressed in monocytes. We developed RNA FISH probes for three such genes, *MSN*, *PGK1* and *CFP* (**Table S1**), respectively encoding moesin, phosphoglycerate kinase 1, and complement factor properdin. Preliminary hybridizations with each probe separately confirmed widespread but strictly mono- allelic transcription of these genes in female human monocytes (**Fig S2A,B**), and we used the pooled probes as a positive marker of the Xa in subsequent RNA FISH experiments. A fraction of female monocytes exhibited two transcriptional foci for *TLR7* (**Fig 2A**) or *TLR8* (**Fig 2C**), denoting bi-allelic expression of both genes. The frequency of nuclei with a positive signal for the Xa probes was 35% in average (Fig S2C), whereas the frequency of nuclei positive for either *TLR7* or *TLR8* primary transcripts was >40% in monocytes (Fig S2D). The percentages of monocyte nuclei with biallelic expression of *TLR7* or *TLR8* among total positive cells for either *TLR7* or *TLR8* signals were 10% (95% CI: 4%-16%) and 17% (95% CI: 8%-27%), respectively. Additionally, we ascribed transcription to the Xi in those cells where a single *TLR7* or *TLR8* transcriptional focus was observed separate from a patent Xa as detected by the Xa probe (**Fig 2B** and **2D)**. By these criteria, we observed escape from XCI for *TLR7*, as expected, but also for *TLR8*, in all the donors of our female study group (n = 6; **S1A Data**). The frequency of *TLR7* transcription on the Xi varied donor to donor between 5% and 32% of cells, with a group mean of 13% (**Fig 2E**). For *TLR8*, transcription on the Xi concerned 10% to 34% of cells, with a group mean of 17% (**Fig 2F).**

**Figure 2.**
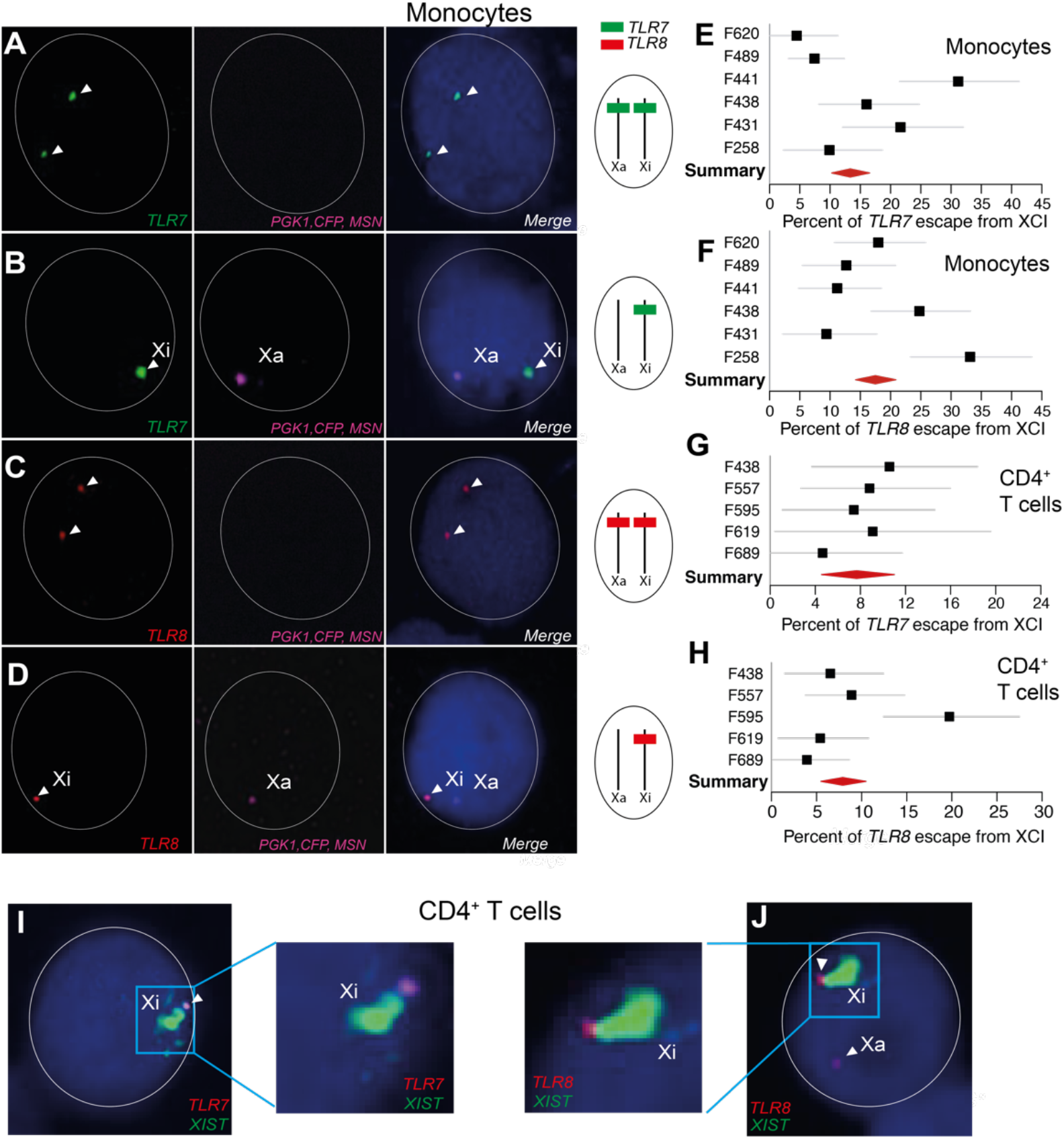
*TLR7* and *TLR8* evade X chromosome inactivation in the monocytes and CD4^+^ T cells of women. (**A-D**) RNA FISH analysis of CD14^+^ monocytes from female donors. The images show confocal microscopy planes of cell nuclei after hybridization with fluorescent probes for transcripts arising from *TLR7* (green), *TLR8* (red), and the marker gene triad transcribed from the Xa only (*PGK1, CFP, MSN*; pink). Nuclei are counterstained with 4ʹ,6- diamidino-2-phenylindole (DAPI; blue). The arrow heads in (**A**, **C**) indicate transcript foci from the two alleles of *TLR7* or *TLR8*. In (**B, D**), the two genes are transcribed only from the Xi (arrowheads). The *TLR7* or *TLR8* hybridization pattern is schematized to the right of each row. (**E-H**) Quantification of the escape from XCI for *TLR7* and *TLR8* from monocytes (**E, F**) and CD4^+^ T cells (**G, H**) The forest plots show the percentage of XCI escape in monocytes or T cells and its 95% confidence interval (CI) in individual female donors (n = 6); the red diamond denotes the meta-analytical group mean and 95% CI.(**I, J**) Confocal microscopy planes of cell nuclei after RNA FISH with *TLR7* (I, pink), *TLR8* (J, red), and *XIST* (green, painting the Xi) probes. Nuclei are counterstained with DAPI (blue). The *TLR7* and *TLR8* transcriptional foci adjacent to the Xi territory confirm that both genes evade XCI.

Because *TLR7* and *TLR8* are also expressed in human CD4^+^ T cells, we investigated whether our findings on XCI escape in CD14^+^ monocytes could be extended to this lymphocyte class. For this, naive CD4^+^ T lymphocytes were stimulated with IL-2, and then activated through CD3 and CD28 (**Fig S3B**). We verified activation on the third day using flow cytometry analysis for CD25 and CD69 levels (**Fig S3C**). As described earlier for monocytes, we concluded to XCI escape in CD4^+^ T cells from women based on the presence of *TLR7* (**Fig S3D**) or *TLR8* (**Fig S3E**) transcripts on both X chromosomes of a cell or, alternatively, on identifying a single transcriptional signal on the Xi (not shown). *TLR7* evaded XCI in 5% to 11% of T cells, with a group mean of 8% (**Fig 2G** and **S1 Data**). For *TLR8*, the frequency of escape cells varied over a 4% to 20% range, with a group mean of 8% (**Fig 2H** and **S1 Data**). On average, XCI escape for these genes was decreased 2- to 3-fold in T cells relative to monocytes. By contrast with monocytes, the XIST hybridization signal characteristic of the Xi could be visualized by RNA FISH in CD4^+^ T cells three days after stimulation through CD3 and CD28 (72). We were thus able to detect *TLR7* and *TLR8* transcripts just next to the inactive Xi territory covered with XIST RNA, further confirming the XCI escape of the two genes (**Fig 2I,J**). Intra-individual levels of XCI escape for *TLR7* and *TLR8* were not significantly correlated in monocytes (Pearson correlation coefficient r = −0.42; p = 0.40; **Fig S4A**) nor in CD4 T cells (r = − 0.06; p = 0.92; **Fig S4C**).

### *TLR7* and *TLR8* evade XCI in monocytes from 47,XXY KS males

In a previous study, we established *TLR7* escape from XCI in monocytes from four 47,XXY men with Klinefelter syndrome (KS) (48). We have studied here five further KS men by RNA FISH. As with female monocytes, we were able to establish XCI escape in all the individuals of this study group based on transcriptional *TLR8* or *TLR7* foci present on both X chromosomes (**Fig 3A**, top and bottom right), or on a single signal ascribable to the Xi (**Fig 3A**, top and bottom left). *TLR7* evaded XCI in 10% to 17% of KS male monocytes (a narrower range than in women), with a group mean of 13% (**Fig 3B** and **S1 Data**). For *TLR8*, the frequency of escape cells varied over an unexpectedly wide range, 8%–49%, with a group mean of 23% (**Fig 3C**). As in females, intra-individual levels of *TLR7* and *TLR8* escape were not correlated (r = −0.03; p = 0.96; **Fig S4B**).

**Figure 3.**
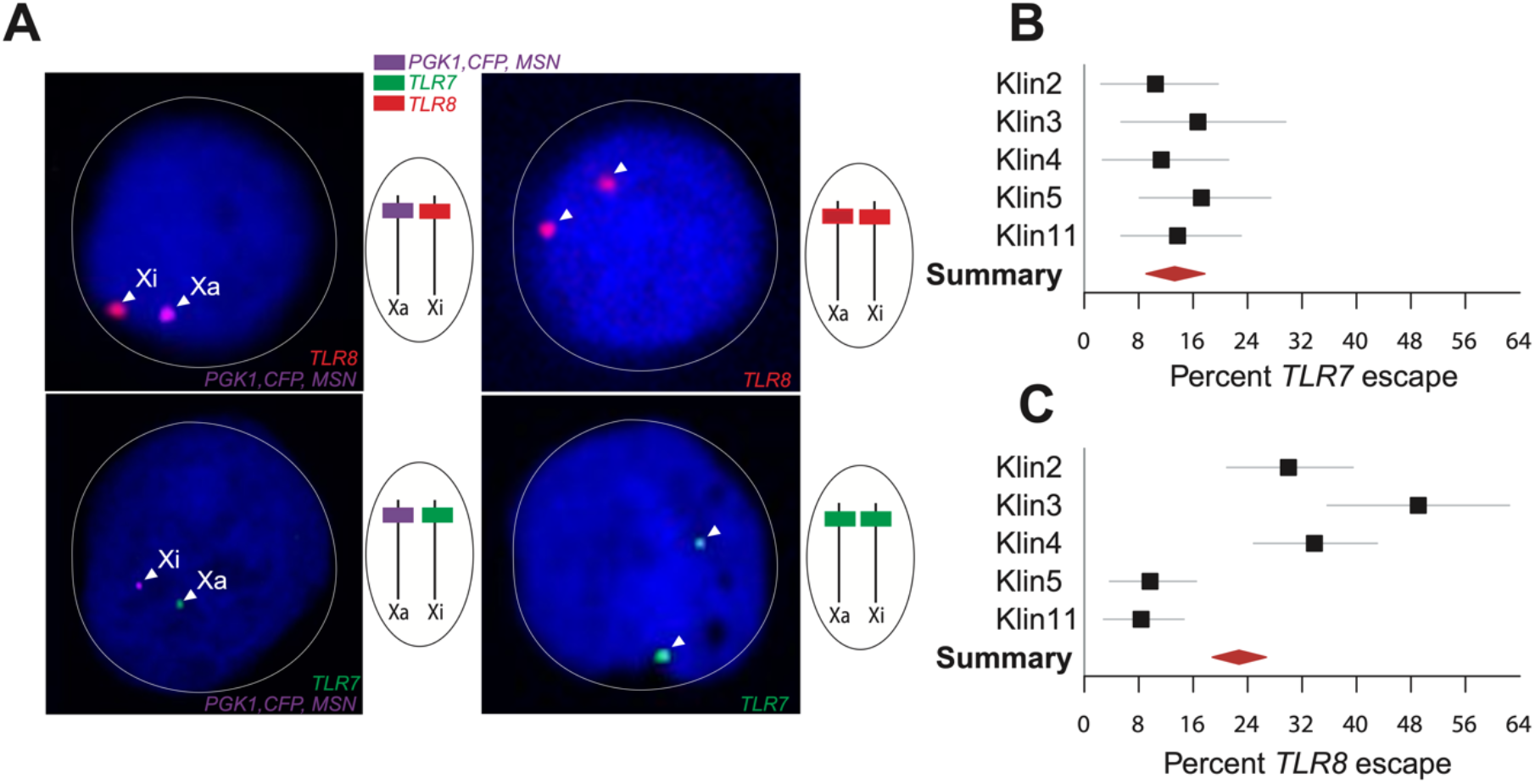
*TLR7* and *TLR8* evade X chromosome inactivation in the monocytes of men with Klinefelter syndrome. (**A**) RNA FISH analysis of CD14^+^ monocytes from men with KS (47,XXY). Confocal microscopy planes of cell nuclei hybridized with fluorescent probes for transcripts arising from *TLR8* (red), *TLR7* (green), and the Xa marker genes (pink). Nuclei are counterstained with DAPI (blue). The arrow heads indicate *TLR8* (top) or *TLR7* (bottom) transcript foci occurring either on both X chromosomes (right) or on the Xi only (left). (**B**, **C**) Quantification of XCI escape for *TLR7* and *TLR8*. The forest plots show the percentage of XCI escape and its 95% CI in KS men (n = 5). The meta-analytical group mean and 95% CI is denoted by the red diamond.

### The frequencies of *TLR7* and *TLR8* transcriptional foci are sex-biased

Regardless of the Xa or Xi chromosome of origin, the proportion of monocytes that exhibited *TLR7* or *TLR8* transcripts exhibited wide inter-individual variation, and was higher overall in the women group (n = 6) than among euploid men (n = 7). There were substantial women-versus-men differences in regard to *TLR7* (**Fig S5A**), with 31% positive cells in women versus only 17% in euploid men, and 18% for men with KS (p < 0.0001). This represents a difference between groups dependent on sex, not on the number of X chromosomes. For *TLR8*, by contrast, between-groups differences were modest (**Fig S5B),** with a female group mean of 30% versus 24% for XY males, and 31% for XXY KS males (n = 5). For women and XY men, intra-group variation in the frequency of *TLR8^+^* cells was distinctly wider than the difference of means between the two groups (**Fig S5B**).

We next determined transcript detection frequencies on the Xa specifically, by considering only those cells positive for the Xa marker probe, and the *TLR7* or *TLR8* foci co- localizing with the Xa signal. Here again, we observed a clear difference between women and XY men, with group means of 48% and 33% of *TLR7* positive cells, respectively (p < 0.0001) (**Fig S5C**). With regard to *TLR8* transcripts, the greatest contrast occurred between the groups of XY and XXY KS men, who exhibited 45% and 61% of positive monocytes, respectively (p < 0.0001), with the women’s group at an intermediate value of 54% (**Fig S5D)**.

The analysis for *TLR7* and *TLR8* transcripts in CD4^+^ T cells, circumscribed to women and XY men, showed a sex bias in the same direction as in monocytes (**Fig S6A** and **S6B, S6 data**). The women’s group mean frequency of *TLR7*-positive cells was 19% versus 15% for XY men. For *TLR8*, the divergence was more marked with 31% of positive cells in women versus 20% in XY men (p < 0.0001). These observations on monocytes and T cells dovetail with our previous observation that, on average, women’s mononuclear blood cells express more TLR7 protein than the cells from normal men (48). The female bias was also clearly visible in the parallel RNA FISH analysis of the transcripts from the Xa specifically (**Fig S6C** and **S6D**), with group means of 48% in women versus 30% in XY men for *TLR7*, and 63% in women versus 39% in XY men for *TLR8*. These sex-dependent divergences were significant for both genes at the p < 0.0001 level.

Overall, the higher counts of *TLR7*- and *TLR8*-positive cells in monocytes and T cells from women relative to the male cells is likely to arise not only from the contribution of escape transcripts of the Xi alleles but also from greater transcriptional frequencies on the Xa in women. This strongly suggests that the single X chromosome of XY men and the Xa of women are functionally non-equivalent as regards the X-linked TLR loci. These observations prompted us to expand our RNA FISH study of *TLR7* and *TLR8* together to quantify the suggested sex bias.

### The combined transcription profile of *TLR7* and *TLR8* is sex-biased in monocytes

Hybridizations combining the *TLR7* and *TLR8* probes hinted at a co-transcriptional sex bias in monocytes, as individual cells from XY men were positive for either *TLR7* (**Fig 4A**) or *TLR8* RNA signals alone (**Fig 4B**), but only rarely for both genes at the same time (**Fig 4C**). By contrast, co-occurring *TLR7* and *TLR8* signals were readily observable in women’s monocytes, where signals from *TLR7*, *TLR8* or both genes occurred in a variety of combinations reflecting the presence of two potential source X chromosomes, Xa and Xi (**Fig 4D–G**). Monocytes from XXY KS males exhibited RNA FISH patterns similar to those of the female cells (**Fig 4G**). In XY men, 95% of signal-positive monocytes, where the two genes are necessarily in *cis*, exhibited transcripts from either *TLR7* (**Fig 4A,G**) or *TLR8* alone (**Fig 4B,G**), and only a consistently small minority (<2% of all cells) transcribed both genes at the same time (**Fig 4C,G** and **Fig 5A**). Among women’s monocytes, *TLR7*^+^ *TLR8*^+^ cell numbers reached 14% on average, a seven-fold increase relative to XY men (**Fig 4D-G** and **Fig 5A**; p < 0.0001). Monocytes from XXY KS men exhibited an intermediate group mean (9% of *TLR7*^+^ *TLR8*^+^ cells **Fig 4G** and **Fig 5A**).

**Figure 4.**
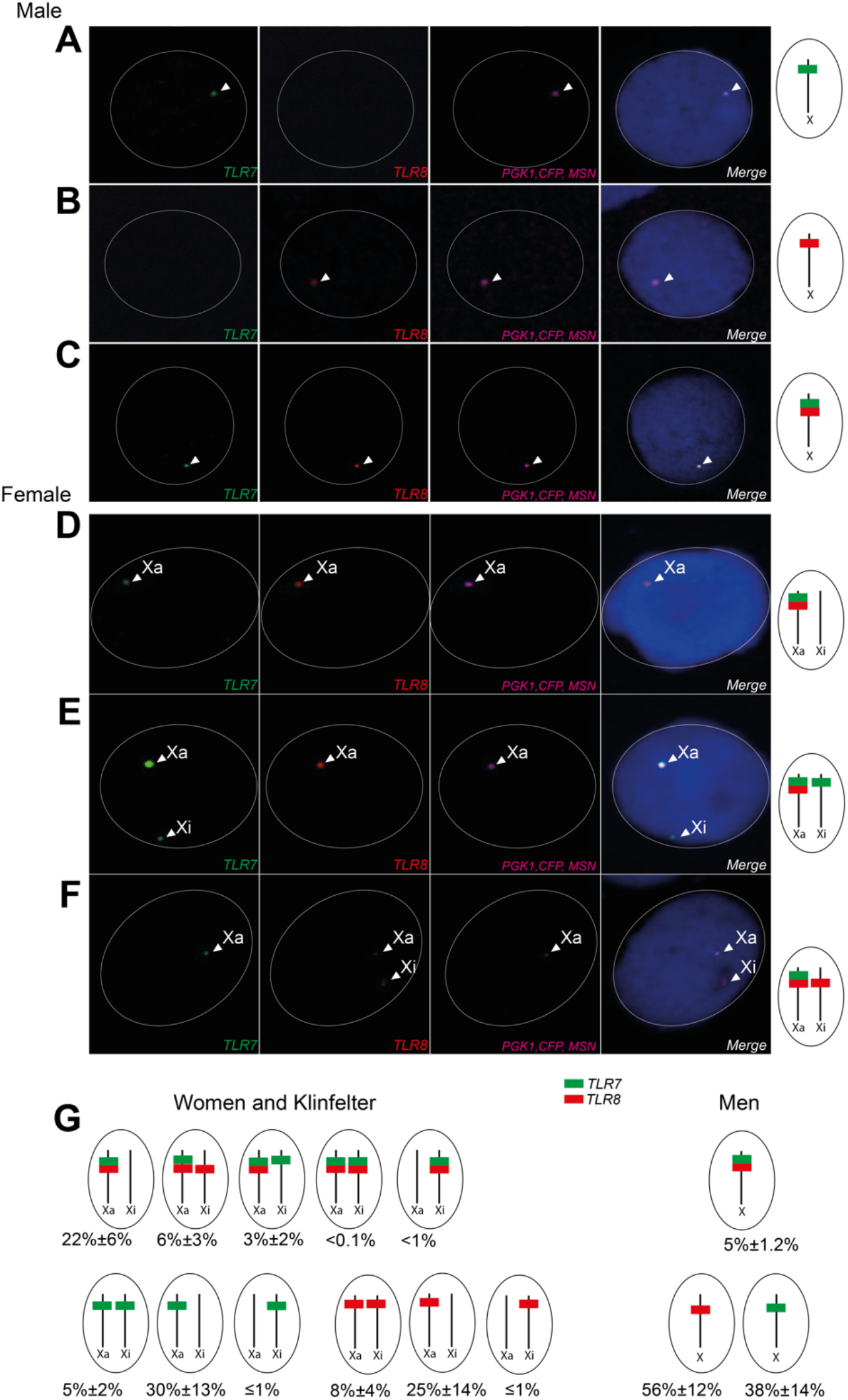
Examples of *TLR7* and *TLR8* combined transcription profiles in euploid male and female monocytes. RNA FISH analysis of CD14^+^ monocytes from male (**A**–**C**) and female (**D**–**F**) donors. Confocal microscopy planes of cell nuclei after RNA FISH for *TLR7* (green), *TLR8* (red), and the Xa marker (pink). Nuclei are counterstained with DAPI (blue). (**A**, **B**) Detection of *TLR7* or *TLR8* transcripts from the single X of male cells. (**C**, **D**) Simultaneous transcription of *TLR7* and *TLR8* on the male X or on the female Xa. (**E, F**) *TLR7* and *TLR8* co-transcription on the female Xa with concomitant *TLR7* (**E**) or *TLR8* (**F**) transcriptional activity on the Xi. (**G**) Schemes of the different patterns of *TLR7* and *TLR8* transcription in our RNA FISH data. The percentages of nuclei (mean ± SD) in females (n=6) and males (n=7) for each RNA FISH profile are shown.

**Figure 5.**
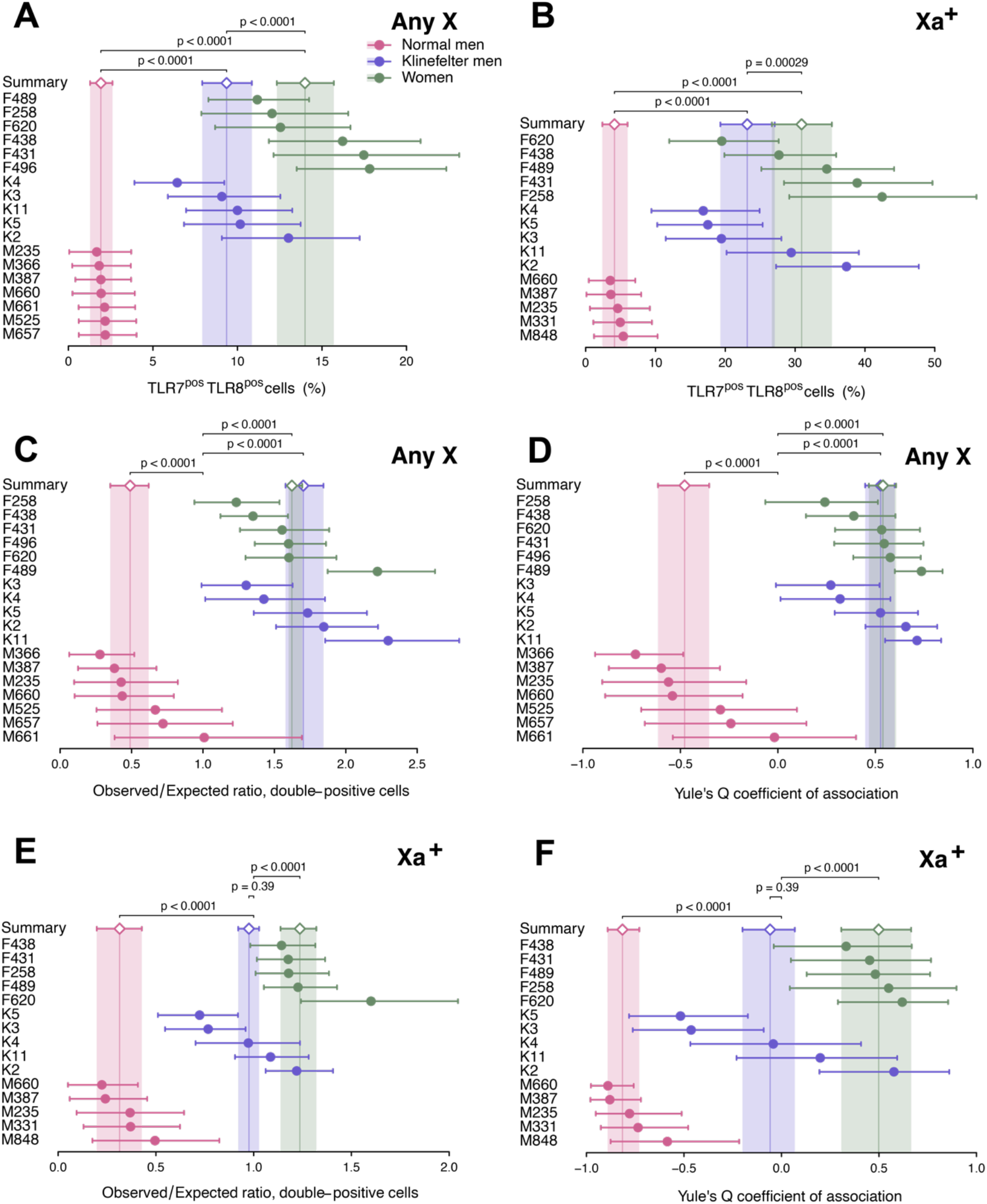
*TLR7* and *TLR8* are transcriptionally non-independent in monocytes. Quantitative analysis of RNA FISH experiments on monocytes from XX women, XY men, and XXY KS men. For each donor, individual cells were scored positive or negative for *TLR7* and for *TLR8* transcripts, and cell counts accordingly cross-classified as a 2×2 contingency table. Descriptive and analytical statistics were computed from each table (**S3 Data**), and summarized group-wise by meta-analysis. The forest plots display the statistics of interest for each donor and its 95% CI (dots and whiskers), and the meta-analytical group means (diamonds) and their 95% CIs (whiskers and shaded areas). In panels (**A, C and E**), cells were scored regardless of the chromosome of origin of the *TLR7* and *TLR8* transcripts (Any X). In (**B, D and F**), only cells positive for the Xa probe (Xa^+^) were counted, and the data is restricted to *TLR7* and *TLR8* signals observed on Xa^+^ male or female X chromosomes. (**A**, **B**) Percentage of cells positive for both *TLR7* and *TLR8* transcripts. (**C**, **D**) Analysis for the *obs*/*exp* ratio of the observed number of double-positive cells to the number of cells in this category expected under the hypothesis of independent transcription of *TLR7* and *TLR8*. (**E**, **F**) Analysis for Yule’s Q coefficient of association in 2×2 tables. The p-values in (**A**, **B**) test the differences between group means; in **C, D, E** and **F**, each p-value tests the divergence of a group summary value relative to the critical value, *obs*/*exp* = 1 or Q = 0; the two-tailed p-values were derived from the group CDs in the meta-analytical summarization. Women and XY men display significant deviations from Q = 0, of opposite signs, signifying transcriptional non-independence of *TLR7* and *TLR8*.

This contrast between women and XY men persisted when we next compared the single X of men with the Xa of women **(Fig 5B).** The frequency of simultaneous transcription of *TLR7* and *TLR8* was again 7-fold greater on the Xa of women (group mean 31%) than on the X of XY men (4.1%; 95% CI: 2.4%–6.0%), while the overall frequency of positive events (for *TLR7*, *TLR8*, or both genes together) was similar between women, normal men and men with Klinefelter syndrome (**Fig S5E**). Reciprocally, the frequency of monogenic expression (either *TLR7* or *TLR8*) was two-fold enriched on the X of XY men (group mean 69%) compared with the Xa of women (38%), with XXY KS men in between the two other groups (53%) (**Fig S5F**). These observations indicate that the Xa of women and XXY KS men is non-equivalent with the single X of XY men regarding the combined transcription of *TLR7* and *TLR8* in monocytes.

### *TLR7* and *TLR8* are transcriptionally non-independent in monocytes

The low frequency of monocytes from XY men where *TLR7* and *TLR8* were transcribed at the same time suggested that the two genes are transcriptionally non-independent from each other. To investigate this possibility, we cross-classified the cell counts in the RNA FISH data as 2×2 contingency tables, i.e., monocytes were stratified depending on the presence of signals for both *TLR7* and *TLR8*, either gene alone, or neither gene (**S3 Data**). This allowed the theoretical (expected) cell counts in each table cell to be calculated under the null hypothesis of independence between the two genes. We used this information to compute the observed-to-expected ratio (*obs*/*exp*) for double-positive events, namely the ratio of observed *TLR7^+^ TLR8^+^* cell counts to the expected number of cells in this stratum assuming transcriptional independence between *TLR7* and *TLR8*. Non-independence would be denoted by a deviation from the critical value, *obs*/*exp* = 1, as a trend for either mutually exclusive (*obs*/*exp* < 1) or co-dependent (*obs*/*exp* > 1) transcription. As shown in **Fig 5C**, monocytes from euploid males comprised only one-half of the expected number of cells displaying *TLR7* and *TLR8* transcripts simultaneously, suggesting mutually exclusive transcription (*obs*/*exp* = 0.49; 95% CI: 0.35– 0.62). Remarkably, we observed an excess of comparable magnitude for double-positive cells among monocytes from females (*obs*/*exp* = 1.62; 95% CI: 1.60–1.70) and KS males (*obs*/*exp* = 1.70; 95% CI: 1.58–1.84), indicating a trend for co-dependent transcription.

In parallel, we used a classical measure of association between two nominal variables, Yule’s Q coefficient of association in 2×2 tables (61). Q can be intuitively interpreted by analogy with a correlation coefficient: Q = −1 would denote mutually exclusive transcription of *TLR7* and *TLR8* in single cells; Q = 0, independent transcription at either locus; and Q = 1, that both genes are always either on or off at the same time. We computed Yule’s Q for each individual, together with the corresponding meta-analytical group summaries for this statistic (**Fig 5D**). Q = −0.48 (95% CI: −0.61 to −0.35) in euploid men; Q = 0.54 (95% CI: 0.47–0.61) in women; and Q = 0.53 (95% CI: 0.45–0.60) in KS men. Deviations from independence (Q = 0) were significant for all groups (p < 0.0001 in all instances). This analysis confirmed the transcriptional non-independence between *TLR7* and *TLR8* expression in women and KS men (Q > 0), and the opposite patterns of transcriptional association in euploid men (Q < 0).

The preceding analyses scored the cells without regard to the chromosome of origin of the transcripts in women and KS men (“Any X” data), but we carried out a parallel scoring procedure (**S4 Data**) restricted to Xa^+^ cells and considering only the alleles on the Xa. This analysis revealed a similar excess of *TLR7-TLR8* co-transcription among women with an elevated *obs*/*exp* ratio (*obs*/*exp* = 1.24; 95% CI: 1.14–1.32) relative to XY men (*obs*/*exp* = 0.31; 95% CI: 0.20–0.43) (**Fig 5E**). The values for Yule’s Q coefficient of association in the Xa- specific data paralleled the patterns for the *obs*/*exp* ratio, and confirmed a negative association between the transcription of *TLR7* and that of *TLR8* in euploid men (Q = −0.82; 95% CI: −0.89 to −0.73), and a positive association in women (Q = 0.50; 95% CI: 0.31–0.66) (**Fig 5F**). Consistent with *obs*/*exp* ≈ 1, the group value for Yule’s Q among KS men was not significantly different from Q = 0 denoting independence, but we concluded that our group of five 47 XXY KS males non-homogeneous, because it encompassed euploid male-like, female-like, and neutral patterns of *TLR7-TLR8* co-transcription on the Xa (**Fig 5F**).

Taken together, these observations indicate the occurrence of mutually exclusive transcription of the genes *TLR7* and *TLR8* in *cis* in the monocytes from euploid men, in parallel with co-dependent transcription of these genes on the Xa of the monocytes from women, and a heterogenous phenotype in 47,XXY men.

### *TLR7-TLR8* transcriptional dependency differs between the monocytic Xa and Xi

Because only a fraction of the female monocytes analyzed exhibited XCI escape, the cross-classified cell counts based only on the Xi signals encompassed fewer positive cells, with instances of zero events in the double-positive (TLR7^+^ TLR8^+^) stratum (**S2 Data**). To analyze these sparse data for Yule’s Q, we pooled the cell counts group-wise to increase statistical power (**S2 Data**). **Fig S7A** shows the pooled data as 2×2 contingency tables of observed frequencies, in a comparison with the frequencies expected under the null hypothesis of independent transcription at either Xi locus. The 95% CIs for Q straddle the critical value, Q = 0 in both the women and KS men groups, and the corresponding p-values from Monte Carlo χ^2^ tests were non-significant (**Fig S7B**). This result points to the absence of transcriptional association between the two genes on the Xi (**Fig S7B**). Next, we contrasted these *TLR7*-*TLR8* co-transcription data for the Xi with the data for the Xa of women and KS men (**Fig 5E,F and S5 Data**). We performed Monte Carlo χ^2^ tests on 3×2 tables of cell counts to formally compare the relative proportions of TLR7^+^ TLR8^+^, TLR7^−^ TLR8^+^, and TLR7^+^ TLR8^−^ cells in the Xa and the Xi of individual donors. The tests demonstrated significant differences between the Xa and the Xi for all individuals (**S5 Data**), and there is therefore a conclusive divergence between the marked *TLR7*-*TLR8* transcriptional co-dependency on the Xa (**Fig 5E,F**) and the trend for non-dependency observed on the Xi alleles (**Fig S7A,B).**

### *TLR7* and *TLR8* are transcriptionally non-independent in CD4^+^ T cells

Further to the study of monocytes, we applied a similar strategy to CD4^+^ T cells from women and euploid men. We scored first the cells regardless of the Xa marking (Any X data), and **Fig 6A** shows that, similar to monocytes, *TLR7*^+^ *TLR8*^+^ events among female T cells were more frequent than among the male cells: 13% of cells versus only 2% in males, a 6:1 ratio consistent with the 7:1 ratio observed earlier in monocytes (**S6 Data**). There was a clear excess of *TLR7*^+^ *TLR8*^+^ cell counts among the female T cells (*obs*/*exp* = 2.27; 95% CI: 2.15–2.40) (**Fig 6B**), a pattern even more pronounced here than in monocytes. The male cells again displayed a shortfall in the expected number of *TLR7*^+^ *TLR8*^+^ events (*obs*/*exp* = 0.77; 95% CI: 0.56–1.00). Yule’s Q coefficient confirmed the strong positive association in women (Q = 0.79; 95% CI: 0.75–0.84) (**Fig 6C**). The parallel analysis restricted to the Xa (i.e., of transcripts for a *TLR7*- *TLR8* gene pair in an obligate *cis* topology) revealed a similar contrast (**Fig 6D**) between the excess of double positive cells in women: *obs*/*exp* = 1.19; 95% CI: 1.15–1.23) and the shortfall in euploid men (*obs*/*exp* = 0.55; 95% CI: 0.42–0.69). On the Xa, the negative association was conclusive (Q = −0.56; 95% CI: −0.68 to −0.43; p < 0.0001) and of a similar magnitude to the association observed earlier in male monocytes (**Fig 5E**). The analysis for Yule’s Q corroborated the positive transcriptional association of the two Xa genes in women’s T cells (**Fig 6F**). In summary, the observations in both CD14^+^ monocytes and CD4^+^ T cells outline a pattern of mutually exclusive transcription for the adjacent *TLR7* and *TLR8* genes in the cells from euploid men, and an opposite pattern of co-dependent transcription of these genes in the cells from women. This is further proof of functional non-equivalence for this locus between the single X chromosome of men and the active X chromosome of women.

**Figure 6.**
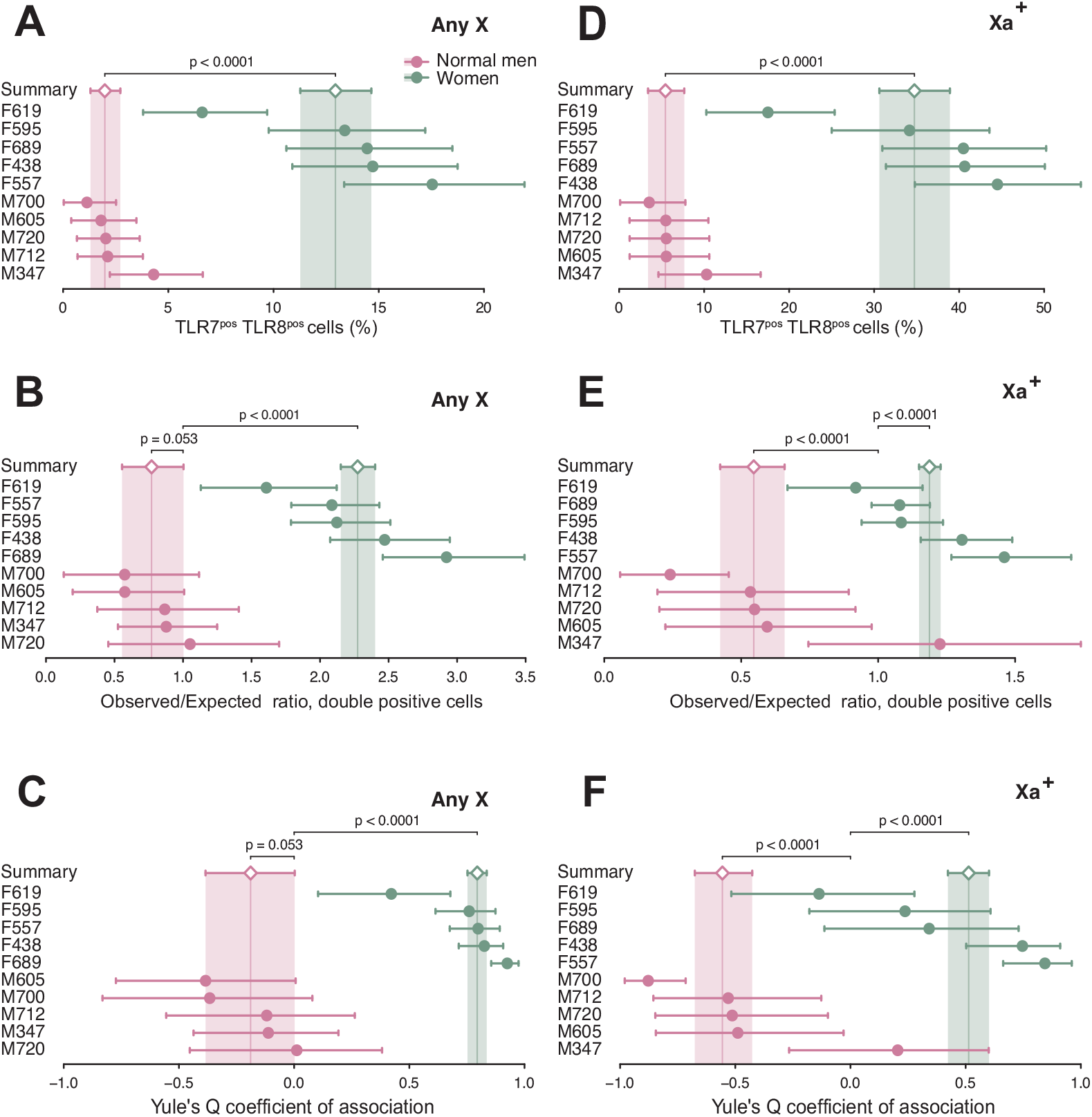
*TLR7* and *TLR8* are transcriptionally non-independent in CD4^+^ T cells. Quantitative analysis of RNA FISH experiments on stimulated CD4^+^ T cells from women and XY men. Cell counts from RNA FISH experiments were processed as in Fig 4. The forest plots display the statistic of interest for each donor and its 95% CI (dots and whiskers), together with the meta-analytical group means (diamonds) and their 95% CIs (whiskers and shaded areas). In (**A**–**C**), T cells were scored regardless of the chromosome of origin of the *TLR7* and *TLR8* transcripts (Any X). In (**D**–**F**), only cells positive for the Xa probe (Xa^+^) were counted, and the data is restricted to *TLR7* and *TLR8* signals observed on those Xa^+^ male or female chromosomes. (**A**, **D**) Percentage of T cells positive for both *TLR7* and *TLR8* transcripts. (**B**, **E**) Analysis for the *obs*/*exp* ratio of the observed number of double-positive cells to the number of cells in this category expected under the hypothesis of independent transcription of *TLR7* and *TLR8*. (**C**, **F**) Analysis for Yule’s Q coefficient of association in 2×2 tables. The p-values in (**A**, **D**) test the differences between group means; in (**B**, **C**, **E**, and **F**), each p-value tests the divergence of a group summary value relative to the critical value, *obs*/*exp* = 1 or Q = 0; the two-tailed p-values were derived from the group CDs in the meta-analytical summarization. Women and euploid men display deviations of opposite signs from Q = 0, signifying transcriptional non-independence of *TLR7* and *TLR8*.

### Female immune cells express higher levels of TLR8 protein than male cells

We previously reported that female PBMCs expressed higher levels of TLR7 protein by western blot, including the full-length (140-kDa) and proteolytically mature (75-kDa) forms of the protein (56, 73). We performed a similar analysis by comparing TLR8 expression between male and female PBMCs using a highly specific TLR8-specific antibody (6). Normalized TLR8 protein expression was significantly higher in female than in male PBMCs (**Fig 7A,B**), despite similar proportions of monocytes between either sex (**Fig 7C**, **Fig S8AB**). Because monocytes were found to express the highest level of TLR8 protein compared with other immune cell populations (not shown), we next analyzed TLR8 expression by flow cytometry within monocyte subsets defined by the expression profile of the CD14 and CD16 markers (**Fig. 7D**, **Fig S8A**). Whereas CD14^+^CD16^−^ classical and CD14^+^CD16^+^ intermediate monocytes all expressed TLR8 protein to variable degrees, only 60% on average of non-classical CD16^+^CD14^−^ monocytes positively stained for TLR8 (**Fig 7D**). Geometric mean fluorescence intensities (GMFIs) of TLR8^+^ cells revealed higher TLR8 protein expression for all subsets of female monocytes than in their male counterparts (**Fig 7E-G**). Together, using two different highly specific mAbs (6, 40), our data provide evidence for higher TLR8 protein expression in female than in male leukocytes, including all monocyte subpopulations.

**Figure 7.**
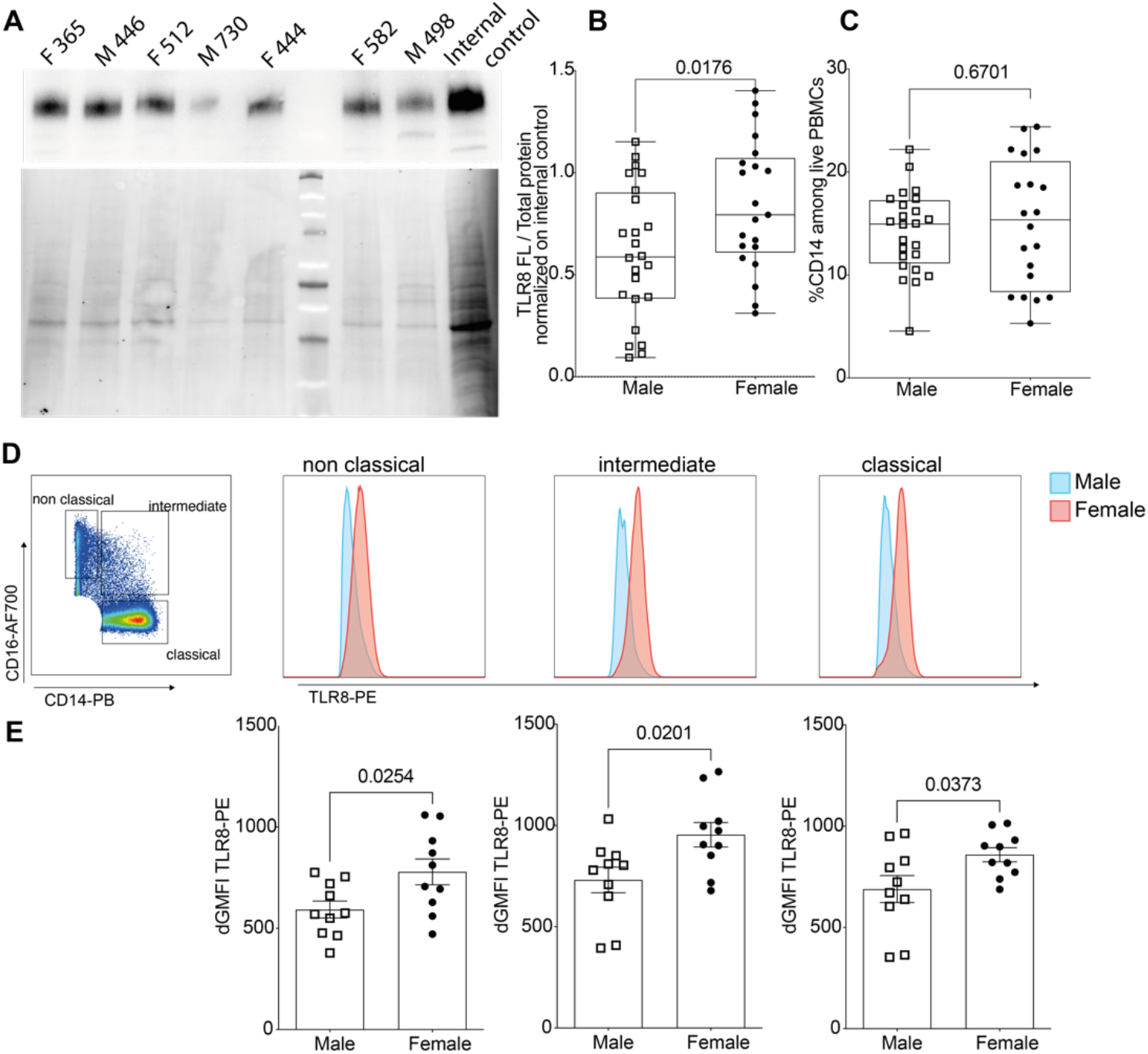
TLR8 protein is expressed at higher level in female than in male immune cells. **(A)** Western blot analysis of TLR8 expression (upper panel) and total protein staining as control (lower panel). (**B**) Densitometric signals from the full-length (120-kDa) form of TLR8 were normalized to total protein and then to an internal standard PBMC lysate that was loaded in each gel for inter-gel data normalization. Results for male (n = 24) and female (n = 21) donors are shown, and data were pooled from four independent experiments. (**C**) Cytometrically- determined frequency of CD14^+^ monocytes among PBMCs from the same donors. (**D**) Classical monocytes were defined as Lin^−^ CD14^+^ CD16^−^, intermediate Lin^−^ CD14^+^ CD16^+^ and non- classical Lin^−^ CD14^−^ CD16^+^ cells as shown in Supplementary **Fig S8A**. TLR8 expression was measured by intracellular co-staining with anti-TLR8-PE and anti-TLR8-APC antibodies as shown in **Fig S8A**. (**D**,**E**) Anti-TLR8-PE staining intensity, gated on double TLR8-positive cells was calculated and expressed as ΔGMFI with background staining subtracted. (**D**) Representative histogram profiles from male and female donors are shown. (**E**) Data from individual male (n = 10) and female (n = 10) donors were pooled from two independent experiments. Statistical differences between groups were analyzed using the Mann and Whitney test; actual p-values are shown.

## Discussion

Here, we conducted a comprehensive single-cell resolution analysis of the transcriptional regulation of the X-linked genes encoding RNA-specific Toll-like receptors, *TLR7* and *TLR8*, in CD14^+^ monocytes and CD4^+^ T lymphocytes, where both receptors are expressed. By analyzing for primary transcript expression relative to the Xa and Xi chromosome territories, we unequivocally demonstrated that *TLR8*, like *TLR7*, escapes XCI in female and in 47,XXY KS male monocytes. We show also that both genes escape XCI in a substantial proportion of T cells despite lower mRNA expression levels than in monocytes (16, 74), with *TLR7* and *TLR8* bi-allelic cells observable in all our female donors. When we analyzed *TLR7* and *TLR8* transcripts together, distinct co-expression profiles emerged between our three study groups. Surprisingly, these differences were attributable not only to the ability of females and KS males to express *TLR7* and *TLR8* on the Xi, but also to the joint transcriptional behavior of the *TLR7*-*TLR8* gene pair on the active X chromosome specifically. Monocytes and T cells from women and KS men exhibited higher-than-expected frequencies of cells co-transcribing the two genes, in contrast to a striking trend against simultaneous transcription of the *TLR7* and *TLR8* genes on the single X of euploid men. Corroborating our RNA FISH results, we found higher TLR8 protein expression in female than in male leukocytes, including all monocyte subpopulations. In summary, our findings provide compelling evidence for sex-specific transcriptional regulation of the X-linked TLR locus on the active X of healthy subjects, which may have important consequences for the functional make-up of monocyte and T-cell populations.

The key findings of the present work are thus the marked association between the transcription of *TLR7* and that of *TLR8* in single cells, and that this association is of positive sign in women and negative in normal men. Simultaneous transcription of *TLR7* and *TLR8* was disproportionally frequent in female monocytes and T cells, and disproportionally scarce in the cells from normal men, and we traced this back to the behavior of the alleles carried in cis on the Xa of women and on the single X chromosome of euploid men. In some of our KS patients, by contrast, there was discordance between the whole-cell and Xa-specific patterns of *TLR7*- *TLR8* transcription, which could have arisen from undetected 46,XY/47,XXY mosaicism (75). Overall, our findings imply that the Xa in XX and XXY karyotypes, and the single X of normal men are functionally non-equivalent as regards the TLR genes. Besides, transcriptional non-independence between *TLR7* and *TLR8* indicates that these genes, which encode the two isoforms of the human TLR for single-stranded RNA, form a co-regulated gene cluster. We propose to name this cluster the X-linked TLR locus to acknowledge this previously unsuspected layer of transcriptional regulation.

Because RNA FISH provides a single-cell snapshot of the transcriptional state of each allele at the time of cell harvest, our data must be interpreted within the notion that genes are not transcribed continuously but in bursts or pulses separated by silent periods (76). Alleles permissive for transcription but analyzed between pulses may thus appear negative, and hybridization-positive cell counts probably underestimate the proportion of expressing cells or the frequency of escape from XCI. This likely explains that, in female monocytes, the average frequency for *TLR7* escape determined here by RNA FISH is lower (13.3% versus 30%) than in our previous study, which employed instead reverse transcription and PCR amplification on the mRNA pool of individual cells (48). Regardless of the technique employed, our results are compatible with the notion that the *TLR7* and *TLR8* genes on the Xi may not be poised for transcription in all cells, and that this status may be relatively stable within cells as previously suggested (48, 77). In favor of this hypothesis, we previously provided evidence for a causal link between X chromosome dosage, XCI escape and increased functional TLR7-driven responses in human B cells (48) and pDCs (78), where bi-allelic *TLR7* expression at single-cell resolution was strongly correlated with enhanced responsiveness to cognate ligands. Along the same line, female pDCs with bi-allelic *TLR7* transcription displayed heightened basal levels of mRNA for IFN-α and IFN-β, which suggests that baseline TLR7 sensing of endogenous ligands contributes to the functional heterogeneity of pDCs (78, 79). This mechanism and its effects should be investigated in further human immune cell populations, such as monocytes and macrophages, in respect of TLR7 but also TLR8. A relevant study in mice has recently identified a robust sexual dimorphism in tissue macrophages (80) by using transcriptome and ATAC (assay for transposase-accessible chromatin) profiling of untreated and IFN-induced immune cells. This study suggested that female macrophages exhibited a higher basal potential of innate immunity pathways prior to immune challenge, and that the resulting enhancement of immune alertness made females less susceptible to infectious diseases (80). Future work should delve deeper into the heterogeneity of these and further immune cell populations, such as T lymphocytes, and possibly define the role of endogenous X-linked TLR signaling in the acquisition of this sex-specific functional heterogeneity.

We hypothesize that the versatile sex-specific transcriptional patterns in the X-linked TLR locus arise from either *cis* or *trans*-acting factors, or both. The significant difference between the Xa and the Xi of individual females regarding *TLR7*-*TLR8* joint transcriptional patterns clearly argues for *cis*-acting factors. The analyses of available Hi-C datasets suggest that although *TLR7* and *TLR8* lie very close to each other on the X chromosome, and fall within the same domain of interactions, the two genes seem to form different sub-domains of interactions that may explain the versatility of the regulation of their expression, with conditions of co-regulation, or co-exclusion or independence, which could be influenced by sex-specific factors, genetic predisposition or the inflammatory environment.

A limitation of the present work is that RNA FISH is not informative on the past or future evolution of the cell, and also that the RNA FISH readout cannot at present be readily correlated with protein quantitation on a single-cell basis, whether by flow cytometry or by emerging techniques such as single-cell proteomics (81). Of note, we previously reported greater levels of TLR7 protein in the PBMCs of women relative to men (48, 56), and the present study shows a similar trend regarding TLR8 protein expression, which positively correlates with the greater frequency of RNA FISH signals observed in female cells. Whether this difference translates into sex-specific responses to TLR8-specific ligands warrants further investigation. Although RNA FISH showed sexual karyotype-biased patterns of transcription at the X-linked TLR locus, this did not translate into specialized cell subpopulations with predominant expression of either TLR7 or TLR8 in males, as the frequencies of TLR8^+^ cells were similar between males and females. Another intriguing question, prompted by evidence of functional attenuation of TLR7 by TLR8 in mouse and human mutants (37, 38, 40), is whether the different patterns of co-transcription observed here determine a sex bias with regard to TLR8:TLR7 protein ratios in individual cells, and hence possibly different outcomes of the exposure to relevant ligands.

### Perspectives and significance

This work constitutes, to our knowledge, an unprecedented single-cell exploration of the transcriptional activity of the X-linked TLR locus according to sex and the sexual karyotype. Together with our earlier study of the *TLR7* gene (48), it provides proof that sex is a critical factor to consider in studying the function and biological significance of the two Toll-like receptors for single-stranded RNA. In addition to the immense worldwide toll of 76 autoimmune disorders (82), and many disabling and life-threatening microbial pathogens, these new insights into the biology of TLR7 and TLR8 are of translational significance to vaccine design, where development of TLR7 and TLR8 agonists as vaccine adjuvants constitutes a promising field of research (83).

## Data Availability

All data produced in the present work are contained in the manuscript

## Abbreviations

CD: confidence distribution
CI: confidence interval
DAPI: 4ʹ,6-diamidino-2-phenylindole
Hi-C: high-resolution chromosome conformation capture analysis
IFN: interferon
KS: Klinefelter syndrome
mAb: monoclonal antibody
obs/exp: observed-to-expected ratio
PBMCs: peripheral blood mononuclear cells
pDCs: plasmacytoid dendritic cells
RNA FISH: RNA fluorescence in situ hybridization
TAD: topologically associating domain
TLR: Toll- like receptor
Xa: active X chromosome
XCI: X chromosome inactivation
Xi: inactive X chromosome

## Acknowledgments

We are grateful to the staff of the core facilities at Infinity, Toulouse, for excellent technical support, especially P.-E. Paulet and R. Romieu-Mourez (immune monitoring biobank); F. L’Faqihi, V. Duplan-Eche, A-L Iscache, H. Garnier, and P. Menut (cytometry and cell sorting facility); and S. Allart, S. Lachambre, and L. Lobjois (cell imaging facility). We also thank M. Savignac (Inserm U1291, Infinity) for helpful discussions and advice, and Dr. Honghai Zhang for insights into TLR molecular evolution. The technical assistance of Flora Abbas (Inserm U1291, Infinity) is also gratefully acknowledged.

## Authors’ contributions

AY, JC, FB, JEM and JCG conceptualized the present study. AY, JC, JEM and JCG analyzed the data and supervised experiments. SG provided essential resources. AY, CC and BFL performed experiments and analyzed the data. JC performed the Hi-C analysis. AY, JEM, FJB and JCG secured funding for this project. AY, JEM and JCG wrote the manuscript with input from their co-authors. All authors read and approved the final version of the manuscript.

## Declarations

### Ethics approval and consent to participate

This study was approved by the relevant ethics board as described in the Materials and Methods section. Written consent was obtained from each patient or, for child participants, from the legal guardian.

### Consent for publication

Not applicable.

### Availability of data and materials

The data supporting the findings of this study are available within the article and its Supplementary Information. R scripts developed for the analysis of RNA FISH data are publicly available in the Zenodo repository, and the relevant identifiers are provided in the Materials and Methods section.

## References

1. Beignon A-S, McKenna K, Skoberne M, Manches O, DaSilva I, Kavanagh DG, et al. Endocytosis of HIV-1 activates plasmacytoid dendritic cells via Toll-like receptor–viral RNA interactions. J Clin Invest. 2005;115(11):3265–75.

2. Diebold SS, Kaisho T, Hemmi H, Akira S, Reis e Sousa C. Innate antiviral responses by means of TLR7-mediated recognition of single-stranded RNA. Science. 2004;303(5663):1529–31.

3. Heil F, Hemmi H, Hochrein H, Ampenberger F, Kirschning C, Akira S, et al. Species-specific recognition of single-stranded RNA via Toll-like receptor 7 and 8. Science. 2004;303(5663):1526–9.

4. Lund JM, Alexopoulou L, Sato A, Karow M, Adams NC, Gale NW, et al. Recognition of single-stranded RNA viruses by Toll-like receptor 7. Proc Natl Acad Sci U S A. 2004;101(15):5598–603.

5. Melchjorsen J, Jensen Søren B, Malmgaard L, Rasmussen Simon B, Weber F, Bowie Andrew G, et al. Activation of innate defense against a paramyxovirus is mediated by RIG-I and TLR7 and TLR8 in a cell-type-specific manner. J Virol. 2005;79(20):12944–51.

6. Greulich W, Wagner M, Gaidt MM, Stafford C, Cheng Y, Linder A, et al. TLR8 Is a Sensor of RNase T2 Degradation Products. Cell. 2019;179(6):1264–75 e13.

7. Ugolini M, Gerhard J, Burkert S, Jensen KJ, Georg P, Ebner F, et al. Recognition of microbial viability via TLR8 drives TFH cell differentiation and vaccine responses. Nat Immunol. 2018;19(4):386–96.

8. Caetano BC, Carmo BB, Melo MB, Cerny A, dos Santos SL, Bartholomeu DC, et al. Requirement of UNC93B1 reveals a critical role for TLR7 in host resistance to primary infection with Trypanosoma cruzi. J Immunol. 2011;187(4):1903–11.

9. Biondo C, Malara A, Costa A, Signorino G, Cardile F, Midiri A, et al. Recognition of fungal RNA by TLR7 has a nonredundant role in host defense against experimental candidiasis. Eur J Immunol. 2012;42(10):2632–43.

10. Barrat FJ, Meeker T, Gregorio J, Chan JH, Uematsu S, Akira S, et al. Nucleic acids of mammalian origin can act as endogenous ligands for Toll-like receptors and may promote systemic lupus erythematosus. J Exp Med. 2005;202(8):1131–9.

11. Shibata T, Ohto U, Nomura S, Kibata K, Motoi Y, Zhang Y, et al. Guanosine and its modified derivatives are endogenous ligands for TLR7. Int Immunol. 2015.

12. Barrat FJ, Elkon KB, Fitzgerald KA. Importance of Nucleic Acid Recognition in Inflammation and Autoimmunity. Annu Rev Med. 2016;67:323–36.

13. Shimizu T. Structural insights into ligand recognition and regulation of nucleic acid-sensing Toll-like receptors. Curr Opin Struct Biol. 2017;47:52–9.

14. Tanji H, Ohto U, Shibata T, Taoka M, Yamauchi Y, Isobe T, et al. Toll-like receptor 8 senses degradation products of single-stranded RNA. Nat Struct Mol Biol. 2015;22(2):109–15.

15. Zhang Z, Ohto U, Shibata T, Krayukhina E, Taoka M, Yamauchi Y, et al. Structural Analysis Reveals that Toll-like Receptor 7 Is a Dual Receptor for Guanosine and Single-Stranded RNA. Immunity. 2016;45(4):737–48.

16. Guiducci C, Gong M, Cepika AM, Xu Z, Tripodo C, Bennett L, et al. RNA recognition by human TLR8 can lead to autoimmune inflammation. J Exp Med. 2013;210(13):2903–19.

17. Hornung V, Barchet W, Schlee M, Hartmann G. RNA recognition via TLR7 and TLR8. In: Bauer S, Hartmann G, editors. Toll-Like Receptors (TLRs) and Innate Immunity. Handbook of Experimental Pharmacology: Springer; 2008. p. 71–86.

18. Dominguez-Villar M, Gautron AS, de Marcken M, Keller MJ, Hafler DA. TLR7 induces anergy in human CD4(+) T cells. Nat Immunol. 2015;16(1):118–28.

19. Fabié A, Mai LT, Dagenais-Lussier X, Hammami A, van Grevenynghe J, Stäger S. IRF- 5 promotes cell death in CD4 T cells during chronic infection. Cell Rep. 2018;24(5):1163–75.

20. Li L, Liu X, Sanders KL, Edwards JL, Ye J, Si F, et al. TLR8-Mediated Metabolic Control of Human Treg Function: A Mechanistic Target for Cancer Immunotherapy. Cell Metab. 2019;29(1):103–23 e5.

21. Peng G, Guo Z, Kiniwa Y, Voo KS, Peng W, Fu T, et al. Toll-like receptor 8-mediated reversal of CD4+ regulatory T cell function. Science. 2005;309(5739):1380–4.

22. Salvi V, Nguyen HO, Sozio F, Schioppa T, Gaudenzi C, Laffranchi M, et al. SARS-CoV-2-associated ssRNAs activate inflammation and immunity via TLR7/8. JCI Insight. 2021;6(18).

23. Laurent P, Yang C, Rendeiro AF, Nilsson-Payant BE, Carrau L, Chandar V, et al. Sensing of SARS-CoV-2 by pDCs and their subsequent production of IFN-I contribute to macrophage-induced cytokine storm during COVID-19. Sci Immunol. 2022;7(75):eadd4906.

24. Barreiro LB, Ben-Ali M, Quach H, Laval G, Patin E, Pickrell JK, et al. Evolutionary dynamics of human Toll-like receptors and their different contributions to host defense. PLoS Genet. 2009;5(7):e1000562.

25. van der Made CI, Simons A, Schuurs-Hoeijmakers J, van den Heuvel G, Mantere T, Kersten S, et al. Presence of genetic variants among young men with severe COVID-19. JAMA. 2020;324(7):663–73.

26. Fallerini C, Daga S, Mantovani S, Benetti E, Picchiotti N, Francisci D, et al. Association of Toll-like receptor 7 variants with life-threatening COVID-19 disease in males: findings from a nested case-control study. eLife. 2021;10:e67569.

27. Kosmicki JA, Horowitz JE, Banerjee N, Lanche R, Marcketta A, Maxwell E, et al. Pan-ancestry exome-wide association analyses of COVID-19 outcomes in 586,157 individuals. Am J Hum Genet. 2021;108(7):1350–5.

28. Solanich X, Vargas-Parra G, van der Made CI, Simons A, Schuurs-Hoeijmakers J, Antolí A, et al. Genetic screening for TLR7 variants in young and previously healthy men with severe COVID-19. Front Immunol. 2021;12.

29. Asano T, Boisson B, Onodi F, Matuozzo D, Moncada-Velez M, Maglorius Renkilaraj MRL, et al. X-linked recessive TLR7 deficiency in ∼1% of men under 60 years old with life-threatening COVID-19. Sci Immunol. 2021;6(62):eabl4348.

30. Lind NA, Rael VE, Pestal K, Liu B, Barton GM. Regulation of the nucleic acid-sensing Toll-like receptors. Nat Rev Immunol. 2022;22(4):224–35.

31. Sharma S, Fitzgerald KA, Cancro MP, Marshak-Rothstein A. Nucleic Acid-Sensing Receptors: Rheostats of Autoimmunity and Autoinflammation. J Immunol. 2015;195(8):3507–12.

32. Pisitkun P, Deane JA, Difilippantonio MJ, Tarasenko T, Satterthwaite AB, Bolland S. Autoreactive B cell responses to RNA-related antigens due to TLR7 gene duplication. Science. 2006;312(5780):1669–72.

33. Deane JA, Pisitkun P, Barrett RS, Feigenbaum L, Town T, Ward JM, et al. Control of toll-like receptor 7 expression is essential to restrict autoimmunity and dendritic cell proliferation. Immunity. 2007;27(5):801–10.

34. Ricker E, Manni M, Flores-Castro D, Jenkins D, Gupta S, Rivera-Correa J, et al. Altered function and differentiation of age-associated B cells contribute to the female bias in lupus mice. Nat Commun. 2021;12(1):4813.

35. Berland R, Fernandez L, Kari E, Han J-H, Lomakin I, Akira S, et al. Toll-like receptor 7-dependent loss of B cell tolerance in pathogenic autoantibody knockin mice. Immunity. 2006;25(3):429–40.

36. Christensen SR, Shupe J, Nickerson K, Kashgarian M, Flavell RA, Shlomchik MJ. Toll- like receptor 7 and TLR9 dictate autoantibody specificity and have opposing inflammatory and regulatory roles in a murine model of lupus. Immunity. 2006;25(3):417–28.

37. Demaria O, Pagni PP, Traub S, de Gassart A, Branzk N, Murphy AJ, et al. TLR8 deficiency leads to autoimmunity in mice. J Clin Invest. 2010;120(10):3651–62.

38. Desnues B, Macedo AB, Roussel-Queval A, Bonnardel J, Henri S, Demaria O, et al. TLR8 on dendritic cells and TLR9 on B cells restrain TLR7-mediated spontaneous autoimmunity in C57BL/6 mice. Proc Natl Acad Sci U S A. 2014;111(4):1497–502.

39. Brown GJ, Cañete PF, Wang H, Medhavy A, Bones J, Roco JA, et al. TLR7 gain-of- function genetic variation causes human lupus. Nature. 2022;605(7909):349–56.

40. Fejtkova M, Sukova M, Hlozkova K, Skvarova Kramarzova K, Rackova M, Jakubec D, et al. TLR8/TLR7 dysregulation due to a novel TLR8 mutation causes severe autoimmune hemolytic anemia and autoinflammation in identical twins. Am J Hematol. 2022;97(3):338–51.

41. Liu G, Zhang H, Zhao C, Zhang H. Evolutionary history of the Toll-like receptor gene family across vertebrates. Genome Biol Evol. 2020;12(1):3615–34.

42. Benton MJ. Vertebrate Palaeontology. 4th ed: Wiley Blackwell; 2015.

43. Dossin F, Heard E. The Molecular and Nuclear Dynamics of X-Chromosome Inactivation. Cold Spring Harb Perspect Biol. 2021.

44. Skuse D, Printzlau F, Wolstencroft J. Sex chromosome aneuploidies. In: Geschwind DH, Paulson HL, Klein C, editors. Handbook of Clinical Neurology. 147: Elsevier; 2018. p. 355–76.

45. Carrel L, Willard HF. X-inactivation profile reveals extensive variability in X-linked gene expression in females. Nature. 2005;434(7031):400–4.

46. Peeters SB, Cotton AM, Brown CJ. Variable escape from X-chromosome inactivation: identifying factors that tip the scales towards expression. Bioessays. 2014;36(8):746–56.

47. Tukiainen T, Villani AC, Yen A, Rivas MA, Marshall JL, Satija R, et al. Landscape of X chromosome inactivation across human tissues. Nature. 2017;550(7675):244–8.

48. Souyris M, Cenac C, Azar P, Daviaud D, Canivet A, Grunenwald S, et al. TLR7 escapes X chromosome inactivation in immune cells. Sci Immunol. 2018;3(19):eaap8855.

49. Abbas F, Cenac C, Youness A, Azar P, Delobel P, Guery JC. HIV-1 infection enhances innate function and TLR7 expression in female plasmacytoid dendritic cells. Life Sci Alliance. 2022;5(10).

50. Scofield RH, Bruner GR, Namjou B, Kimberly RP, Ramsey-Goldman R, Petri M, et al. Klinefelter’s syndrome (47,XXY) in male systemic lupus erythematosus patients: support for the notion of a gene-dose effect from the X chromosome. Arthritis Rheum. 2008;58(8):2511–7.

51. Harris VM, Sharma R, Cavett J, Kurien BT, Liu K, Koelsch KA, et al. Klinefelter’s syndrome (47,XXY) is in excess among men with Sjögren’s syndrome. Clin Immunol. 2016;168:25–9.

52. Scofield RH, Lewis VM, Cavitt J, Kurien BT, Assassi S, Martin J, et al. 47XXY and 47XXX in Scleroderma and Myositis. ACR Open Rheumatol. 2022.

53. Caron G, Duluc D, Fremaux I, Jeannin P, David C, Gascan H, et al. Direct stimulation of human T cells via TLR5 and TLR7/8: flagellin and R-848 up-regulate proliferation and IFN- gamma production by memory CD4+ T cells. J Immunol. 2005;175(3):1551–7.

54. de Marcken M, Dhaliwal K, Danielsen AC, Gautron AS, Dominguez-Villar M. TLR7 and TLR8 activate distinct pathways in monocytes during RNA virus infection. Sci Signal. 2019;12(605):eaaw1347.

55. Meas HZ, Haug M, Beckwith MS, Louet C, Ryan L, Hu Z, et al. Sensing of HIV-1 by TLR8 activates human T cells and reverses latency. Nature communications. 2020;11(1):147.

56. Azar P, Mejia JE, Cenac C, Shaiykova A, Youness A, Laffont S, et al. TLR7 dosage polymorphism shapes interferogenesis and HIV-1 acute viremia in women. JCI Insight. 2020;5(12):e136047.

57. Smit A, Hubley R, Green P. RepeatMasker Open-4.0 2013-2015 [Available from: https://www.repeatmasker.org.

58. Greenfield A, Carrel L, Pennisi D, Philippe C, Quaderi N, Siggers P, et al. The UTX gene escapes X inactivation in mice and humans. Hum Mol Genet. 1998;7(4):737–42.

59. Chaumeil J, Augui S, Chow JC, Heard E. Combined immunofluorescence, RNA fluorescent in situ hybridization, and DNA fluorescent in situ hybridization to study chromatin changes, transcriptional activity, nuclear organization, and X-chromosome inactivation. In: Hancock R, editor. The Nucleus. 1: Nuclei and subnuclear components. Totowa, N.J.: Humana Press; 2008. p. 297–308.

60. R Core Team. R: A language and environment for statistical computing. R Foundation for Statistical Computing, Vienna, Austria. 2022.

61. Berry KJ, Johnston JE, Mielke PW. Fourfold Contingency Tables, I. The Measurement of Association: A Permutation Statistical Approach. Cham, Switzerland: Springer; 2018. p. 511–76.

62. Yang G, Cheng JQ, Xie M, Qian W. *gmeta*: Meta-analysis via a unified framework of confidence distribution. R package version 2.3–1. 2021.

63. Xie Mg, Singh K. Confidence distribution, the frequentist distribution estimator of a parameter: A review. Int Stat Rev. 2013;81(1):3–39.

64. Durand NC, Robinson JT, Shamim MS, Machol I, Mesirov JP, Lander ES, et al. Juicebox provides a visualization system for Hi-C contact maps with unlimited zoom. Cell Syst. 2016;3(1):99–101.

65. Rao SSP, Huntley MH, Durand NC, Stamenova EK, Bochkov ID, Robinson JT, et al. A 3D map of the human genome at kilobase resolution reveals principles of chromatin looping. Cell. 2014;159(7):1665–80.

66. Phanstiel DH, Van Bortle K, Spacek D, Hess GT, Shamim MS, Machol I, et al. Static and dynamic DNA loops form AP-1-bound activation hubs during macrophage development. Mol Cell. 2017;67(6):1037–48.e6.

67. Zhang X, Jeong M, Huang X, Wang XQ, Wang X, Zhou W, et al. Large DNA methylation nadirs anchor chromatin loops maintaining hematopoietic stem cell identity. Mol Cell. 2020;78(3):506–21.e6.

68. Lieberman-Aiden E, van Berkum NL, Williams L, Imakaev M, Ragoczy T, Telling A, et al. Comprehensive mapping of long-range interactions reveals folding principles of the human genome. Science. 2009;326(5950):289–93.

69. Giorgetti L, Lajoie BR, Carter AC, Attia M, Zhan Y, Xu J, et al. Structural organization of the inactive X chromosome in the mouse. Nature. 2016;535(7613):575–9.

70. Savarese F, Flahndorfer K, Jaenisch R, Busslinger M, Wutz A. Hematopoietic precursor cells transiently reestablish permissiveness for X inactivation. Mol Cell Biol. 2006;26(19):7167–77.

71. Syrett CM, Sindhava V, Hodawadekar S, Myles A, Liang G, Zhang Y, et al. Loss of Xist RNA from the inactive X during B cell development is restored in a dynamic YY1- dependent two-step process in activated B cells. PLoS Genet. 2017;13(10):e1007050.

72. Wang J, Syrett CM, Kramer MC, Basu A, Atchison ML, Anguera MC. Unusual maintenance of X chromosome inactivation predisposes female lymphocytes for increased expression from the inactive X. Proc Natl Acad Sci U S A. 2016;113(14):E2029–38.

73. Souyris M, Cenac C, Azar P, Daviaud D, Canivet A, Grunenwald S, et al. TLR7 escapes X chromosome inactivation in immune cells. Sci Immunol. 2018;3(19):eaap8855.

74. Hornung V, Rothenfusser S, Britsch S, Krug A, Jahrsdorfer B, Giese T, et al. Quantitative Expression of Toll-Like Receptor 1-10 mRNA in Cellular Subsets of Human Peripheral Blood Mononuclear Cells and Sensitivity to CpG Oligodeoxynucleotides. The Journal of Immunology. 2002;168(9):4531–7.

75. Lanfranco F, Kamischke A, Zitzmann M, Nieschlag E. Klinefelter’s syndrome. Lancet. 2004;364(9430):273–83.

76. Wang YL, Ni TF, Wang W, Liu F. Gene transcription in bursting: a unified mode for realizing accuracy and stochasticity. Biol Rev Camb Philos Soc. 2019;94(1):248–58.

77. Souyris M, Mejía JE, Chaumeil J, Guéry J-C. Female predisposition to TLR7-driven autoimmunity: gene dosage and the escape from X chromosome inactivation. Semin Immunopathol. 2019;41(2):153–64.

78. Hagen SH, Henseling F, Hennesen J, Savel H, Delahaye S, Richert L, et al. Heterogeneous escape from X chromosome inactivation results in sex differences in type I IFN responses at the single human pDC level. Cell Rep. 2020;33(10):108485.

79. Wimmers F, Subedi N, van Buuringen N, Heister D, Vivie J, Beeren-Reinieren I, et al. Single-cell analysis reveals that stochasticity and paracrine signaling control interferon-alpha production by plasmacytoid dendritic cells. Nature communications. 2018;9(1):3317.

80. Gal-Oz ST, Maier B, Yoshida H, Seddu K, Elbaz N, Czysz C, et al. ImmGen report: sexual dimorphism in the immune system transcriptome. Nature communications. 2019;10(1):4295.

81. Marx V. A dream of single-cell proteomics. Nat Methods. 2019;16(9):809–12.

82. Bender M, Christiansen J, Quick M. The terrible toll of 76 autoimmune diseases. Sci Am. 2021;325(3):31–3.

83. Dowling DJ. Recent advances in the discovery and delivery of TLR7/8 agonists as vaccine adjuvants. Immunohorizons. 2018;2(6):185–97.

