## Supplementary Figure S1-S8 for "*TLR8* escapes X chromosome inactivation in human monocytes and CD4^+^ T cells"

### Supplementary Figures

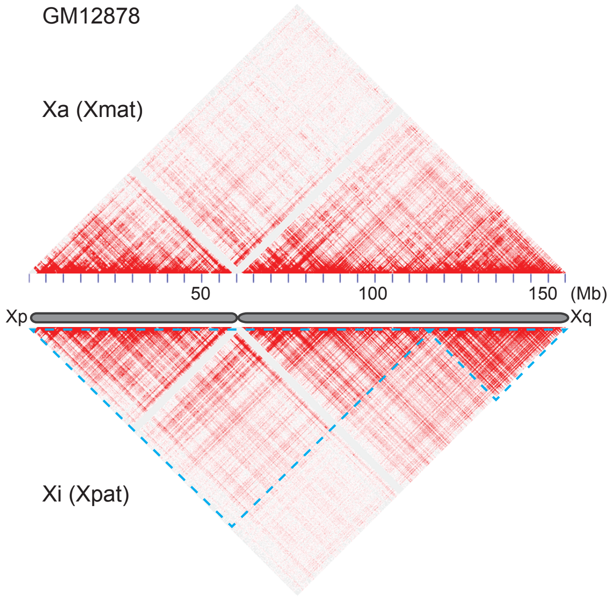


**Figure S1: 3D conformation of the active and inactive X chromosomes in GM12878 female cells.** HiC maps of interactions of the entire maternal active X chromosome (Xa, top panel) or paternal inactive X chromosome (Xi, bottom panel). The Xa is organized into the typical succession of Topologically Associating Domains (TADs), as all the other autosomes, while the Xi lacks most of the TADs and is instead uniquely organized into two mega-domains (blue dashed triangles). Datasets from Rao et al (2014) visualized with the Juicebox software.

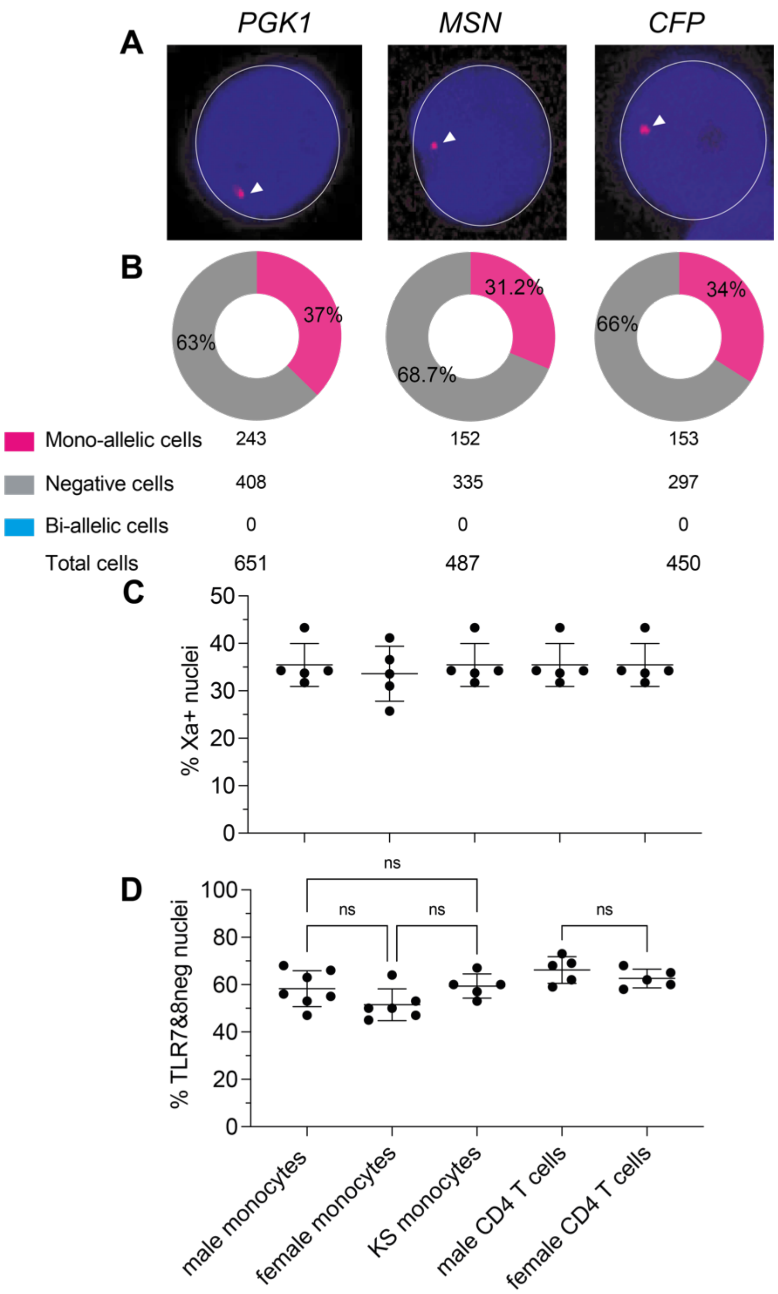


**Figure S2. *PGK1, CFP and MSN* do not escape from X chromosome inactivation.** (**A**)The images show confocal microscopy planes of CD14$^{+}$ monocyte nuclei after hybridization with individual RNA FISH fluorescent probes for *PGK1*, *CFP* and *MSN* transcripts (pink). Nuclei are counterstained with DAPI. (**B**) Frequencies of nuclei showing one pinpoints of the indicated primary transcripts for *PGK1*, *CFP* and *MSN* genes. Nuclei with two pinpoints were never detected indicating that these genes do not escape from XCI. (**C**) Frequencies of Xa+ nuclei positive for the mixed *PGK1*, *CFP* and *MSN*-specific probes in monocytes or CD4+ T cells from the males, females or KS subjects used in the present study. Frequency of nuclei without signals for *TLR7* or *TLR8* are indicated for all conditions. Statistical differences between group were assessed by one-way ANOVA followed by a Tukey's multiple comparisons test. ($p > 0.05$, ns).

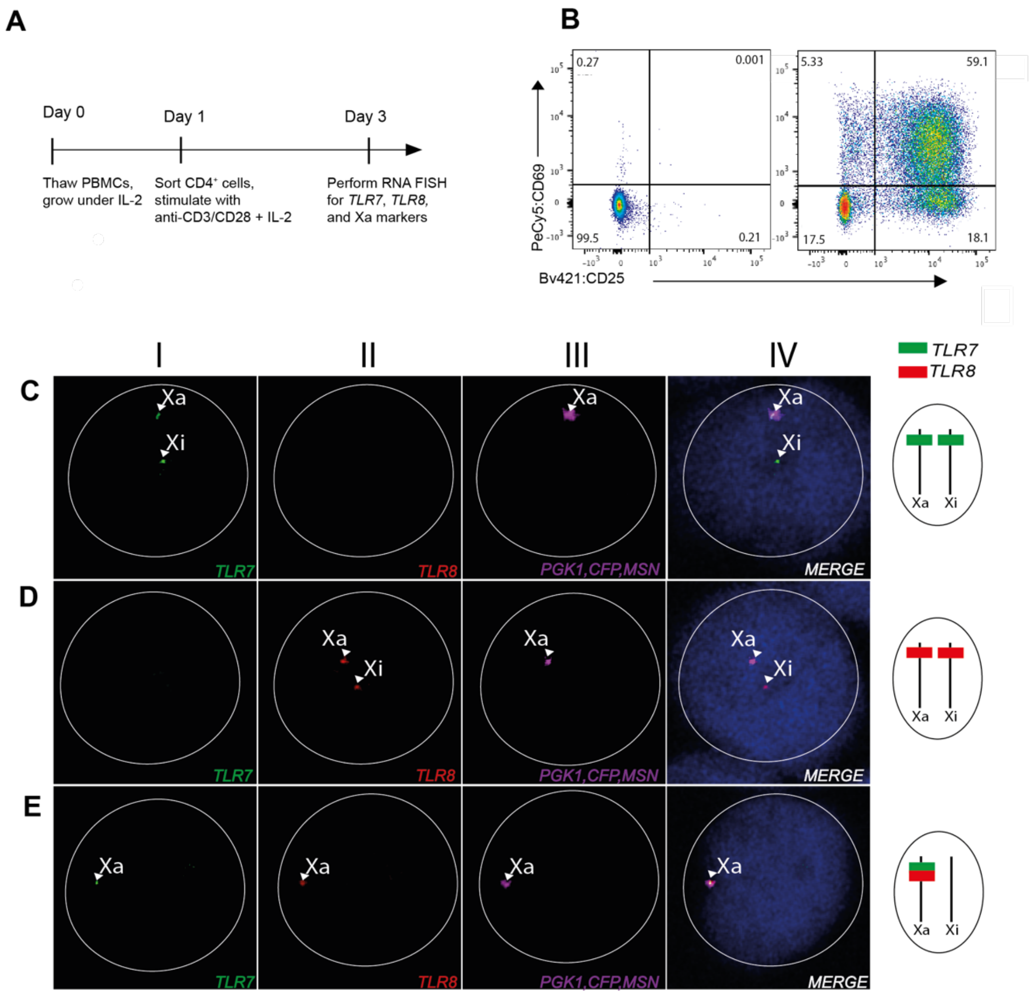


**Figure S3. TLR7 and TLR8 escape from XCI in CD4 T cells.** (**A**) Timeline of the strategy used to isolate and stimulate naive CD4$^+$ T cells from women prior to RNA FISH analysis. (**B**) Flow cytometry scatterplots of CD4$^+$ T cells based on CD25 and CD69 expression. Left, non-activated control cells; right, cells activated with anti-CD3/CD28 microbeads. (**C**–**E**) RNA FISH analysis of T cells. Confocal microscopy planes of female cell nuclei hybridized with fluorescent probes for *TLR8* (red), *TLR7* (green), and Xa marker (pink) transcripts; nuclei are counterstained with DAPI (blue). Arrow heads mark duplicate transcript foci from the two alleles of *TLR7* (**C**, I) or *TLR8* (**D,** II), or a single signal from the allele carried by the Xa (**E**, I and II). The *TLR7* or *TLR8* hybridization pattern is schematized to the right of each row.

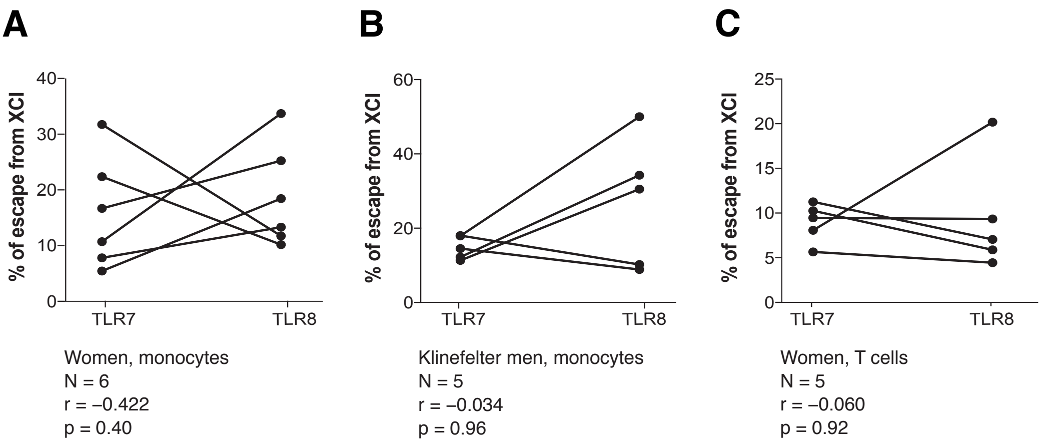


**Figure S4: Absence of correlation between *TLR7* and *TLR8* regarding inter-individual variations in the frequencies of XCI escape.** The graphs show correlation analyses comparing *TLR7* and *TLR8* as to the frequencies of XCI escape in monocytes from women (A), men with Klinefelter syndrome (B) and T cells from women (C).

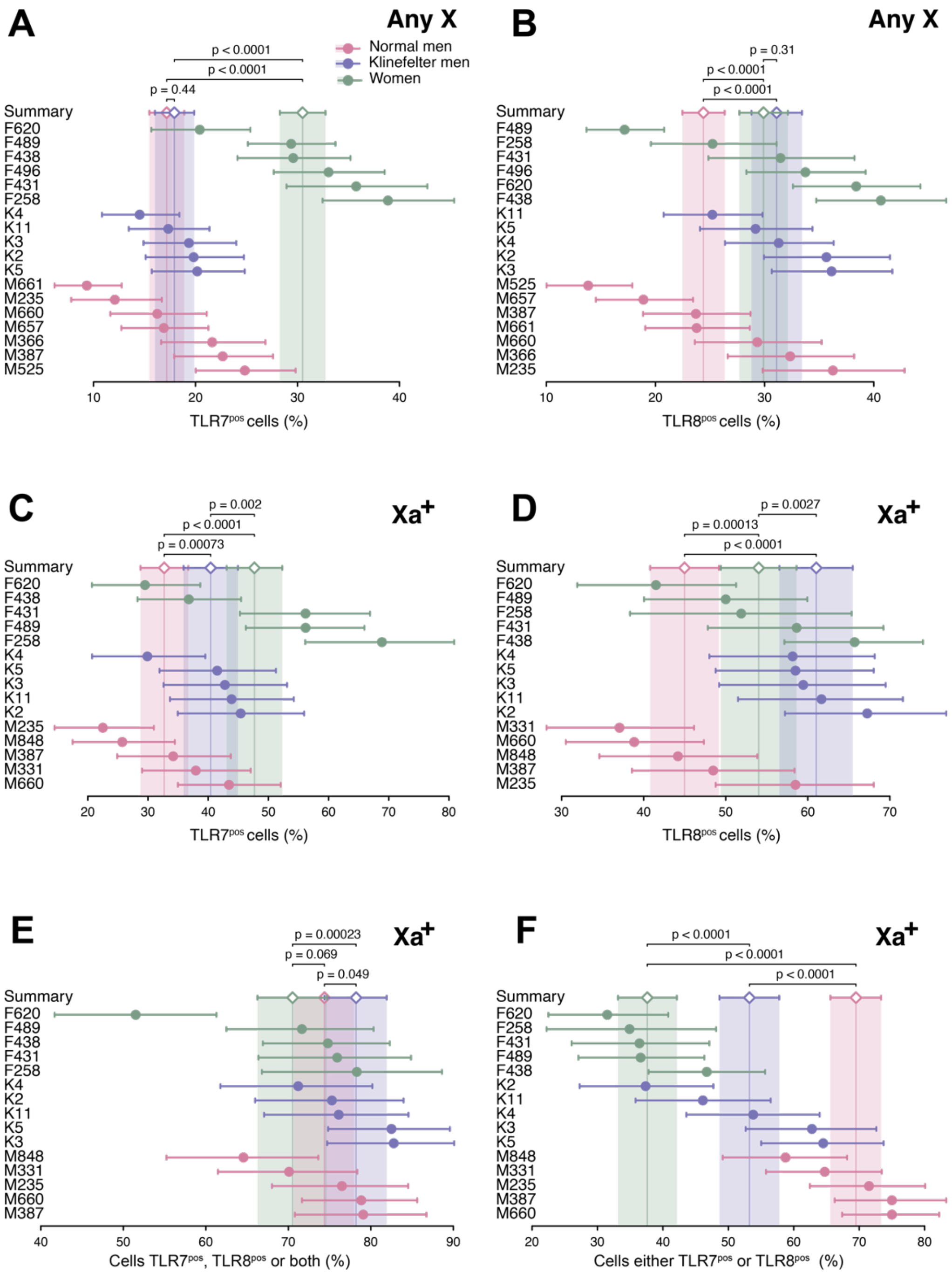


**Figure S5. The frequencies of *TLR7* and *TLR8* transcripts are biased in monocytes from women, normal men, and men with Klinefelter syndrome.** The forest plots display the frequencies of positive cells for the gene or genes of interest in each donor, and its 95% CI (dots and whiskers), together with the meta-analytical group means (diamonds) and their 95% CIs (whiskers and shaded areas). (A, B) Proportion of monocytes positive for *TLR7* (A) or *TLR8* (B) relative to total cells, i.e., regardless of the Xa or Xi chromosome of origin (Any X data) of the RNA FISH signals. (C–F) Proportions relative to the monocyte subset positive for the Xa marker probe, considering only the signals originating from the Xa (Xa$^+$ data).

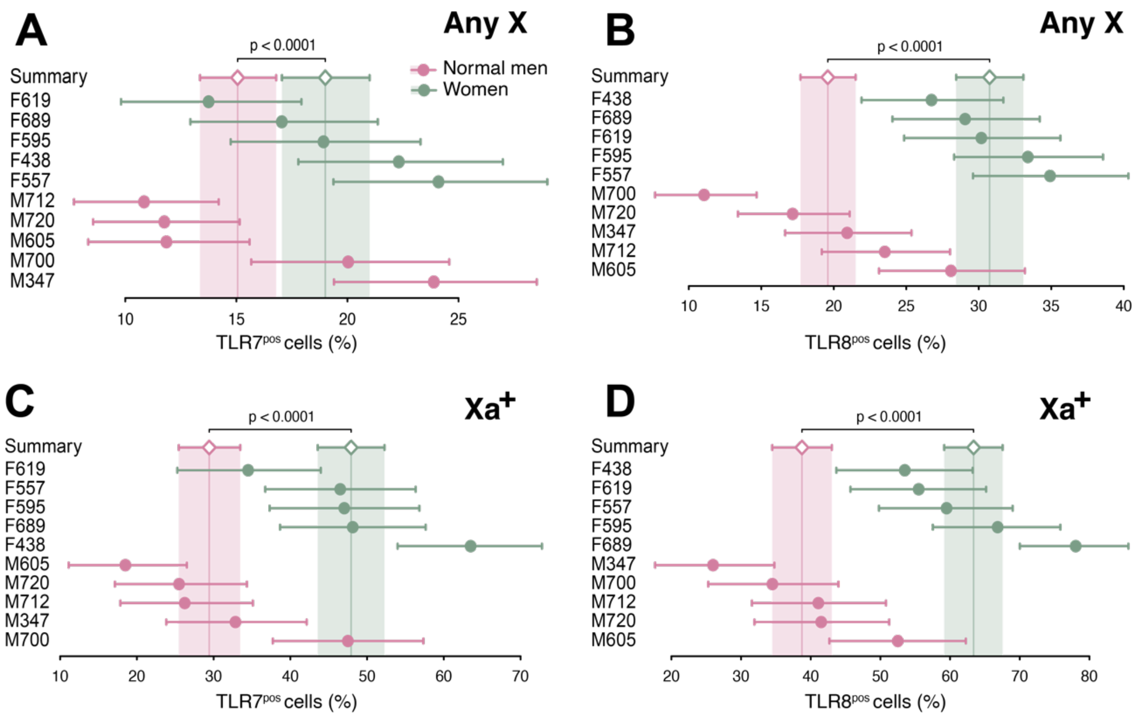


**Figure S6: The frequencies of *TLR7* and *TLR8* transcripts are sex- biased in T cells from women and normal men.** The forest plots display the frequencies of positive cells for the gene or genes of interest in each donor, and its 95% CI (dots and whiskers), together with the meta-analytical group means (diamonds) and their 95% CIs (whiskers and shaded areas). (**A**, **C**) Proportion of activated $CD4^+$ T cells positive for *TLR7* (**A**), *TLR8* (**B**), relative to total cells, i.e., regardless of the Xa or Xi chromosome of origin (Any X data) of the RNA FISH signals. (**C, D**) Proportions relative to the subset of activated $CD4^+$ T cells positive for the Xa marker probe, considering only the signals originating from the Xa ($Xa^+$ data). Cells positive for *TLR7* alone (**C**), *TLR8* alone (**D**), are analyzed separately.

**A**

Observed frequencies

| | | TLR7+ | TLR7- |
|---|---|---|---|
| Women N=6 | TLR8+ | 4 | 99 |
| | TLR8- | 54 | 766 |
| Klinefelter men N=5 | TLR8+ | 3 | 109 |
| | TLR8- | 38 | 546 |

Expected frequencies

| | | TLR7+ | TLR7- |
|---|---|---|---|
| Women N=6 | TLR8+ | 6.5 | 96.5 |
| | TLR8- | 51.5 | 768.5 |
| Klinefelter men N=5 | TLR8+ | 6.6 | 105.4 |
| | TLR8- | 34.4 | 549.6 |

Cell counts according to Xi signals only

**B**

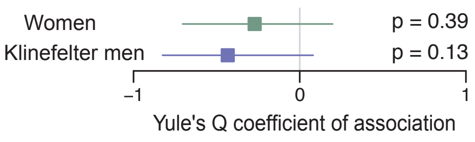


**Figure S7: TLR7 and TLR8 are transcriptionally non-independent on the Xi in monocytes from females or KS males. (A, B)** Analysis of *TLR7* and *TLR8* simultaneous expression from the Xi in monocytes from females and KS males. Single-cell counts, aggregated by donor group, were cross-classified into 2×2 contingency tables according to the two-fold criterium of *TLR7* and *TLR8* transcription on the Xi (**A**, left). The tables on the right show the expected frequencies of cells in each category, derived from the observed frequencies under the assumption that *TLR7* and *TLR8* are mutually independent with regard to transcription on the Xi. (**B**) Forest plot of Yule's Q coefficients of association. Deviations from the critical value that denotes non-association, Q = 0, were not significant. Horizontal whiskers represent 95% CIs; p-values from Monte Carlo $\chi^2$ tests.

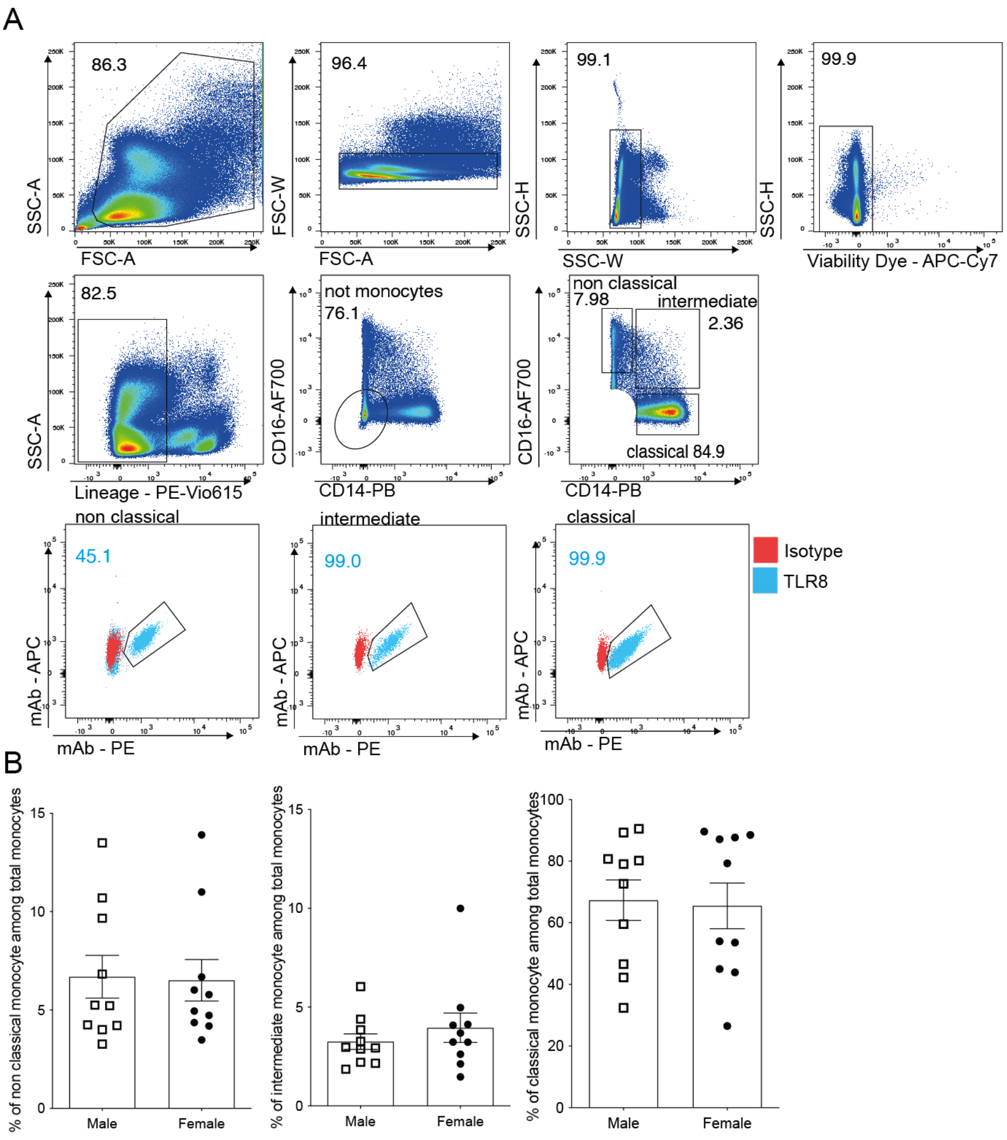


**Figure S8: Gating strategy for intracellular analysis of TLR8 expression in male and female monocyte subsets.** Freshly thawed PBMCs from male or female donors were extracellularly stained with PE-Vio615 Lin- specific antibodies directed to CD3, CD19, CD56, with anti-CD14-PB and anti-CD16-AF700 antibodies, after staining with Viability Dye. Cells were then fixed and permeabilized for staining with anti-TLR8-PE and anti-TLR8-APC antibody, or with PE- or AP-labeled isotype control antibodies. (**A**) The gating strategy used to define monocyte subsets among living mononucleated Lin− (CD3, CD19, CD56) cells is shown. Classical monocytes were defined as Lin− CD14+ CD16−, intermediate Lin− CD14+ CD16+ and non-classical Lin− CD14− CD16+ cells. TLR8 expression was measured by intracellular co-staining with anti-TLR8-PE and anti-TLR8-APC antibodies. The percentage of each monocyte subtype was analyzed to compare male and female and shown in (**B**). No statistical differences were observed between male and female using the Mann and Whitney test.
