## Supplementary tables: S1 Table, S1-S6 Data for "*TLR8* escapes X chromosome inactivation in human monocytes and CD4^+^ T cells"

#### S1 Table. PCR primer pairs used in the preparation of the RNA FISH probes.

##### A. Human *XIST* probe[1].

| Primer pair | Sequences, 5′–3′ | Amplimer size |
|---|---|---|
| XIST-1F + XIST-1R | AATGCTGGTAAAGCCCACAC<br>TCTGGGGACAAGAACCATTC | 516 bp |
| XIST-2F + XIST-2R | GACACCATGGCTACCTGTGA<br>TCCCATTACCCTTTGGATTG | 492 bp |
| XIST-3F + XIST-3R | CAACGAGGAAGCAGGAGTCT<br>CATTTGCTCCTGATCTGTGC | 491 bp |
| XIST-4F + XIST-4R | ATGGAATGGGCAAAGTGGT<br>CGTCTGGGATCCTATGCACT | 477 bp |
| XIST-5F + XIST-5R | AGTGCATAGGATCCCAGACG<br>TTGCAGTTGTCAATGGTCCT | 511 bp |
| XIST-6F + XIST-6R | GTGTCACCAACCATGCTGTC<br>GAGTAGAATCCCGCTTCCTTG | 506 bp |
| XIST-7F + XIST-7R | TTCATAGTCCCAGGGAAAGGT<br>AGGTCCCTGCATCATCTTTG | 508 bp |
| XIST-8F + XIST-8R | GGTGTGAGCAGTTGGGATCT<br>TGCAATTGTCCAGAGTCCTG | 475 bp |
| XIST-9F + XIST-9R | GGGAGGTTCAGACCACACAG<br>GCTGTGTGCTTTTCGTGTTG | 509 bp |
| XIST-10F + XIST-10R | GGTTGGGTTATGCAGCAATC<br>TTGTTGTATCGGGAGGCAGT | 525 bp |
| XIST-11F + XIST-11R | AACACTGCGACAGAACTGGA<br>TTGTGGGTTGTTGCACTCTC | 501 bp |
| XIST-12F + XIST-12R | GACTTCCTCTGCCTGACCTG<br>GGATACCTGCTGATTCCCTTC | 514 bp |

[1]Primer set from our previous publication, Souyris et al., Sci Immunol. 2018; 3:eaap8855.

**B. Human *MSN* probe.**

| Primer pair | Sequences, 5′–3′ | Amplimer size |
|---|---|---|
| MSN-1F + MSN-1R | GTCTGCAGTTGCCTGTCTGT<br>GGCTCAGTCGGCTTGTAGAG | 516 bp |
| MSN-2F + MSN-2R | CTTAGGGGCCAGGGTAACTC<br>ACCTTATTCGGGCAGGAAAT | 508 bp |
| MSN-3F + MSN-3R | TCCTCAAGCTGGTTCGTCTC<br>AGCAGGAATTGCCATGATCT | 529 bp |
| MSN-4F + MSN-4R | CCTTCCCTGAATACCAAGCA<br>TGCCCTCTATGGGTCTGACT | 575 bp |
| MSN-5F + MSN-5R | AGATTTTGGTCACCCCATGA<br>TGAAACACGCACAGAGGAAG | 571bp |
| MSN-6F + MSN-6R | GGAGGTGACTGCCACTTAGC<br>GCAGTTTGCACACAAATCCA | 546 bp |
| MSN-7F + MSN-7R | TCATAACTCCAGGGCAAACA<br>GCAATGCTTTGGACACTCAA | 533 bp |
| MSN-8F + MSN-8R | TGGGATGTGCTGTGCTAATC<br>TGATCCCATGCTCCTGTTG | 515 bp |
| MSN-9F + MSN-9R | GCCACATTATGCTGTGAGAGG<br>CTGCATTTCAAGTGGCATTC | 522 bp |
| MSN-10F + MSN-10R | TTCAAACTGGACCCCAAGTC<br>TGGCTAGCAGGGACTCATCT | 577 bp |
| MSN-11F + MSN-11R | GTGTTGTGGGGAGTGGTTG<br>GATTGCATATGGGCTGGTCT | 523 bp |
| MSN-12F + MSN-12R | AGATCAGGGGAAAGGCAATC<br>TTCCCTCTCCAAGCCTAACA | 513 bp |
| MSN-13F + MSN-13R | GGCAACAGCCTGTCTTAGGT<br>CAGTTCAGTGATGCTTTCCA | 523 bp |
| MSN-14F + MSN-14R | ATTTGGGAAAGGGTGGAGAT<br>TTAATGGAGGCCAAGGCTAA | 595 bp |
| MSN-15F + MSN-15R | GTGGGCCTGGTGAAAATCTA<br>CAAATGCTGAAAGCCCTAGC | 561bp |
| MSN-16F + MSN-16R | GAAGACCCTTGGATTGGTCA<br>CCTTGTTTCACAAGCAAGCA | 530 bp |
| MSN-17F + MSN-17R | CCAGGTGGTAGGAAAGTGGA<br>GCCTGGCCTACAATAAATGC | 584 bp |
| MSN-18F + MSN-18R | GGGTGAAGTGGTTAGGTTGC<br>TTGGAAAATGGGCAGCTATC | 510 bp |
| MSN-19F + MSN-19R | TTGTTCTGTGCGGAACTGAG<br>AACCCCACATGCTTTGACAT | 516 bp |
| MSN-20F + MSN-20R | GTTGGCCACTAATGGAGGTC<br>TGACTATCTGCAAGGCCTCA | 584 bp |
| MSN-21F + MSN-21R | TGGGGAGTGCTCATTAAGCTA<br>TGGACTGTGGAAACACAAGC | 504 bp |
| MSN-22F + MSN-22R | CTTGCCTTCTTGGCTTTGAG<br>CTTCAGGCTCTGGTGACTCTG | 596 bp |

**B. Human *MSN* probe.**

| Primer pair | Sequences, 5′–3′ | Amplimer size |
|---|---|---|
| MSN-23F + MSN-23R | GGTTTGGATCAGCTCCAGTG<br>CACCACCCATTCTAGGGAGA | 566 bp |
| MSN-24F + MSN-24R | GGTAGGTGAGGGGAAACCAT<br>GGATGGGGTAGCATTTCAAG | 566 bp |
| MSN-25F + MSN-25R | TGCATGAGTACCCATTGAATC<br>TGCAAATTTCCATGGAGTGA | 584 bp |
| MSN-26F + MSN-26R | AGATGCCGACATCAGTTCTTG<br>GGGGATACCAATCCTCTCTTG | 596 bp |
| MSN-27F + MSN-27R | AGGAAGTGACCCAATGATGG<br>CTGCCTGCTGTGGGAAGTAT | 584 bp |

**C. Human *PGK1* probe.**

| Primer pair | Sequences, 5′–3′ | Amplimer size |
|---|---|---|
| PGK1-1F + PGK1-1R | AGCCGACTTGTTCTCTCGTC<br>GCCCAAGAGTCCAACGATAC | 570 bp |
| PGK1-2F + PGK1-2R | AGGTCATTTTACTTTCCCTTCC<br>AAACACCACTCTTGGATCCTGT | 624 bp |
| PGK1-3F + PGK1-3R | GGTTCAAATGAGGGCTTGTG<br>TTAAGCAGCTCCTCCAATCC | 608 bp |
| PGK1-4F + PGK1-4R | GGAAGTTACATGGTTCCCTGA<br>AACCATGTCCATCAATGCTTC | 641 bp |
| PGK1-5F + PGK1-5R | GGCAAGAGCAGTTCCTGTGT<br>CCCCTTCCTCTCTAAGGCTA | 638 bp |
| PGK1-6F + PGK1-6R | AATAATCATGGCAGGCACAA<br>CACAGGAACTAGCCCTGAAGA | 559 bp |
| PGK1-7F + PGK1-7R | TGAACTGACAACCAGAATTCAA<br>ATTTTGAACTTGGGACTGACTC | 528 bp |
| PGK1-8F + PGK1-8R | TTCCAATCACTAGTGGCAGATG<br>GACTCAAGCTGCTGGCTACC | 608 bp |
| PGK1-9F + PGK1-9R | TGTTGTCATGTTTCCATCTGC<br>ATCCCCTAGCTTGGAAAGTG | 536 bp |
| PGK1-10F + PGK1-10R | GGTGCTCATTTAGCAATTTTGA<br>GGGGAATGTGCAAAATAGATGT | 559 bp |
| PGK1-11F + PGK1-11R | GGGGAATGTGCAAAATAGATGT<br>GATGAGCTGGATCTTGTCTGC | 506 bp |

**D. Human *CFP* probe.**

| Primer pair | Sequences, 5′–3′ | Amplimer size |
|---|---|---|
| CFP-1F + CFP-1R | GGGCGGATACTGACTCTAGC<br>TCATAGGGCAAACAGCACAG | 546 bp |
| CFP-2F + CFP-2R | GACTGCACACTCGCTTTCAC<br>GTAGCCCTCGGCATATAGCA | 504 bp |
| CFP-3F + CFP-3R | CTAGCTTCTTTGCCCCTCCT<br>TCATAGGGCAAACAGCACAG | 502 bp |
| CFP-4F + CFP-4R | TGCCTTGTCCTGACATTCAA<br>TCAACCTAGGTCCAGGAAGC | 532 bp |
| CFP-5F + CFP-5R | TCCGTTCACAGTCTTGATGC<br>TATTACAGGCGGGAGAGTCG | 553 bp |
| CFP-6F + CFP-6R | ACTCGGCAAGGCAGATACC<br>ATCCGACGGAACATGAAGTC | 539 bp |
| CFP-7F + CFP-7R | CAAAGCAGGGAATCAGCAA<br>GCTCTTGATTGAGGGCAGAG | 555 bp |
| CFP-8F + CFP-8R | AATCGTGAACCCTGAGATGC<br>ACCCCACGAACCTAAGGAGA | 523 bp |
| CFP-9F + CFP-9R | CTCCTCACCAGGACAGCACT<br>TGGTGATCTGGTGGGTTTCT | 502 bp |
| CFP-10F + CFP-10R | CTAGAAACCAAGGGCTGCTG<br>ACTGGAAGCATTGCAGGTG | 587 bp |
| CFP-11F + CFP-11R | TTGAGAAACCCTGAGGTATGG<br>CTGCCAGCCACAGGTGAG | 534 bp |

**E. Human *TLR8* probe.**

| Primer pair | Sequences, 5′–3′ | Amplimer size |
|---|---|---|
| TLR8-1F + TLR8-1R | TCGAACTTTAAGCCTCCAACA<br>TCGAACTTTAAGCCTCCAACA | 534 bp |
| TLR8-2F + TLR8-2R | TAAAGCGGCTTGCTATGATCT<br>ACGGACCACCTGCTATGTCT | 480 bp |
| TLR8-3F + TLR8-3R | ACGGACCACCTGCTATGTCT<br>GGCTTGTCCTATGGTCTTGAGT | 484 bp |
| TLR8-4F + TLR8-4R | CATTGTGCTGAAGCAGCTAAGT<br>CTGAAATTGGATGCTGATGG | 495 bp |
| TLR8-5F + TLR8-5R | TTCCAGGAAGACAGCTTTACG<br>CCTCTCCCTGCCAAATACAG | 471 bp |
| TLR8-6F + TLR8-6R | GGCATGGTCACACAGCACT<br>GCCTCAACATCCGATTTCAA | 488 bp |
| TLR8-7F + TLR8-7R | TCTTGCCCTTCCCATTTATGT<br>GGCCCAAGGAGAATATGTCAG | 487 bp |
| TLR8-8F + TLR8-8R | ACATTAAGCAGCAACATCACGA<br>AGTAACACAGGAGAGTGCCTCA | 487 bp |
| TLR8-9F + TLR8-9R | TGGGCAATAATGATTGAGCTT<br>AATCCTCGTGCCTGAACTGT | 535 bp |
| TLR8-10F + TLR8-10R | AAAACACCTGGCAAATACTGC<br>CTCTGGCAAACCAGAGGGTA | 494 bp |
| TLR8-11F + TLR8-11R | ATATCACAGACGGGGCATTC<br>TTAATTGAAGCACCACCATCA | 487 bp |
| TLR8-12F + TLR8-12R | CAAACTGCCAAGCTCCCTAC<br>TCAGGGGCTGGAAATCATC | 518 bp |
| TLR8-13F + TLR8-13R | AAGGGGAGTTATCCACAGCA<br>TGACATGAGGAATGGCTGAA | 550 bp |
| TLR8-14F + TLR8-14R | GCCACAATGTGCTGCTTATG<br>TTCATTTGGGATGTGCTTCA | 544 bp |
| TLR8-15F + TLR8-15R | GGAAAGCAAGTCCCTGGTAGA<br>TCAAAGGGGTTTCCGTGTAG | 510 bp |
| TLR8-16F + TLR8-16R | AACAATCAACAAATCCGCACT<br>CGGCTCTCTTCAAGGTGGTA | 480 bp |
| TLR8-17F + TLR8-17R | GATGTCATTTGTGCCAGTCCT<br>TAGCCTCTGCAAAGCCAAGTA | 504 bp |

**F. Human *TLR7* probe[1].**

| Primer pair | Sequences, 5′–3′ | Amplimer size |
|---|---|---|
| TLR7-1F + TLR7-1R | TTATAAGGAAGTGGGGAGGAGA<br>TCTTGCCAGCTGGTTTCAG | 477 bp |
| TLR7-2F + TLR7-2R | TGCTCTGCTCTCTTCAACCAG<br>TTTTCTTTGACAGCACTCAAGC | 513 bp |
| TLR7-3F + TLR7-3R | AGAGCCCTATGGTCTTTGGTG<br>CTGGAAACCTTGCCCTCA | 487 bp |
| TLR7-4F + TLR7-4R | TCAGCAACTTCCCTTATTCACA<br>GGATTTCCTGCTCTCCTGACT | 503 bp |
| TLR7-5F + TLR7-5R | GCTTGGAGGAACAGCTGAGT<br>AGACAGGACATATGGGAGCAG | 482 bp |
| TLR7-6F + TLR7-6R | TCAGTTGGTAGCCCTGTAGCA<br>TCAGGCCTAAAGGTAACACTGC | 525 bp |
| TLR7-7F + TLR7-7R | ACGTGGTGCTAACCCCTAGAA<br>TGCTGTCAGGGAAATCTATGGT | 525 bp |
| TLR7-8F + TLR7-8R | TTCTCCCTCACCTGCTTGA<br>AGGAATTTTTCCCTGAAGATCA | 476 bp |
| TLR7-9F + TLR7-9R | TTTCACAGCCTCCATTTTCC<br>TGCCAGTGGGTACAGGAAG | 536 bp |
| TLR7-10F + TLR7-10R | GTGTACCAGGGCAGGTTTAGC<br>ACCATCTAGCCCCAAGGAGT | 513 bp |
| TLR7-11F + TLR7-11R | ACTCCTTGGGGCTAGATGGT<br>CAGTTTTGGCCCAGGTAGAG | 480 bp |
| TLR7-12F + TLR7-12R | CCTCCCGCCTAGCTTACAG<br>GGGCACATGCTGAAGAGAGT | 484 bp |
| TLR7-13F + TLR7-13R | AATGCTTTTGATGCGCTGA<br>TTGAGCAGAAGCCAACTTCA | 490 bp |
| TLR7-14F + TLR7-14R | AGTGAAGTTGGCTTCTGCTCA<br>TGGTGGAGGAAGAGATGTCA | 520 bp |
| TLR7-15F + TLR7-15R | ATCTCTTCCTCCACCAGCAG<br>GGACATTTTCTGGGAAGCTG | 493 bp |
| TLR7-16F + TLR7-16R | CAGCTTCCCAGAAAATGTCC<br>TGGTAACCAGTCCCTTTCCTC | 511 bp |
| TLR7-17F + TLR7-17R | CAAACTGGAAGACCCAAGAGA<br>CAAAACACGCTTTTGGTGTG | 502 bp |

[1]Primer set from our previous publication, Souyris et al., Sci Immunol. 2018; 3:eaap8855.

### S1 Data

##### TLR7 escape from XCI (monocytes, women)

| Donor ID | Total cell count | Escape cell count | Percent escape | 95% CI lower | 95% CI upper |
|---|---|---|---|---|---|
| F620 | 55 | 3 | 5.5 | 0 | 11.3 |
| F489 | 128 | 10 | 7.8 | 3.1 | 12.4 |
| F441 | 85 | 27 | 31.8 | 21.5 | 41.3 |
| F438 | 78 | 13 | 16.7 | 8.1 | 24.7 |
| F431 | 67 | 15 | 22.4 | 12.1 | 32.1 |
| F258 | 56 | 6 | 10.7 | 2.3 | 18.6 |
| Summary | | | 13.3 | 10.3 | 16.6 |

##### TLR8 escape from XCI (monocytes, women)

| Donor ID | Total cell count | Escape cell count | Percent escape | 95% CI lower | 95% CI upper |
|---|---|---|---|---|---|
| F620 | 103 | 19 | 18.4 | 10.8 | 25.8 |
| F489 | 75 | 10 | 13.3 | 5.5 | 20.9 |
| F441 | 85 | 10 | 11.8 | 4.8 | 18.5 |
| F438 | 107 | 27 | 25.2 | 16.7 | 33.2 |
| F431 | 59 | 6 | 10.2 | 2.2 | 17.7 |
| F258 | 86 | 29 | 33.7 | 23.3 | 43.3 |
| Summary | | | 17.5 | 14.2 | 20.9 |

##### TLR7 escape from XCI (monocytes, Klinefelter syndrome men)

| Donor ID | Total cell count | Escape cell count | Percent escape | 95% CI lower | 95% CI upper |
|---|---|---|---|---|---|
| K2 | 53 | 6 | 11.3 | 2.5 | 19.7 |
| K3 | 39 | 7 | 17.9 | 5.4 | 29.6 |
| K4 | 49 | 6 | 12.2 | 2.7 | 21.2 |
| K5 | 61 | 11 | 18.0 | 8.1 | 27.4 |
| K11 | 62 | 9 | 14.5 | 5.4 | 23 |
| Summary | | | 13.3 | 9.1 | 17.7 |

##### TLR8 escape from XCI (monocytes, Klinefelter syndrome men)

| Donor ID | Total cell count | Escape cell count | Percent escape | 95% CI lower | 95% CI upper |
|---|---|---|---|---|---|
| K2 | 95 | 29 | 30.5 | 20.9 | 39.5 |
| K3 | 54 | 27 | 50.0 | 35.7 | 62.5 |
| K4 | 105 | 36 | 34.3 | 24.9 | 43 |
| K5 | 88 | 9 | 10.2 | 3.7 | 16.5 |
| K11 | 90 | 8 | 8.9 | 2.8 | 14.6 |
| Summary | | | 22.7 | 18.8 | 26.7 |

##### TLR7 escape from XCI (CD4+ T cells, women)

| Donor ID | Total cell count | Escape cell count | Percent escape | 95% CI lower | 95% CI upper |
|---|---|---|---|---|---|
| F438 | 71 | 8 | 11.3 | 3.7 | 18.4 |
| F557 | 74 | 7 | 9.5 | 2.7 | 16 |
| F595 | 62 | 5 | 8.1 | 1.1 | 14.6 |
| F619 | 39 | 4 | 10.3 | 0.4 | 19.6 |
| F689 | 53 | 3 | 5.7 | 0 | 11.7 |
| Summary | | | 7.7 | 4.5 | 11 |

##### TLR8 escape from XCI (CD4+ T cells, women)

| Donor ID | Total cell count | Escape cell count | Percent escape | 95% CI lower | 95% CI upper |
|---|---|---|---|---|---|
| F438 | 85 | 6 | 7.1 | 1.5 | 12.4 |
| F557 | 107 | 10 | 9.3 | 3.8 | 14.8 |
| F595 | 109 | 22 | 20.2 | 12.4 | 27.5 |
| F619 | 85 | 5 | 5.9 | 0.7 | 10.8 |
| F689 | 90 | 4 | 4.4 | 0.1 | 8.6 |
| Summary | | | 7.9 | 5.5 | 10.5 |

**S2 Data. RNA FISH analysis of CD14⁺ monocytes, signals on the Xi.** Left, observed frequencies: cell counts cross-classified as 2×2 contingency tables depending on the presence of transcriptional foci for *TLR7* or *TLR8* on the Xi of monocytes from women and from men with Klinefelter syndrome. Right, the corresponding expected frequencies, computed under the null hypothesis of mutually independent transcription of the two genes in *cis* on the Xi. Donor IDs in red.

#### A. Women

| Observed frequencies | | | Expected frequencies | | |
|---|---|---|---|---|---|
| **F620** | TLR7 + | TLR7 – | | TLR7 + | TLR7 – |
| TLR8 + | 0 | 19 | TLR8 + | 0.4 | 18.6 |
| TLR8 – | 3 | 136 | TLR8 – | 2.6 | 136.4 |
| **F489** | TLR7 + | TLR7 – | | TLR7 + | TLR7 – |
| TLR8 + | 1 | 10 | TLR8 + | 0.4 | 10.6 |
| TLR8 – | 6 | 186 | TLR8 – | 6.6 | 185.4 |
| **F438** | TLR7 + | TLR7 – | | TLR7 + | TLR7 – |
| TLR8 + | 1 | 26 | TLR8 + | 1.9 | 25.1 |
| TLR8 – | 12 | 146 | TLR8 – | 11.1 | 146.9 |
| **F431** | TLR7 + | TLR7 – | | TLR7 + | TLR7 – |
| TLR8 + | 1 | 5 | TLR8 + | 0.7 | 5.3 |
| TLR8 – | 14 | 106 | TLR8 – | 14.3 | 105.7 |
| **F296** | TLR7 + | TLR7 – | | TLR7 + | TLR7 – |
| TLR8 + | 1 | 10 | TLR8 + | 1.4 | 9.6 |
| TLR8 – | 13 | 85 | TLR8 – | 12.6 | 85.4 |
| **F258** | TLR7 + | TLR7 – | | TLR7 + | TLR7 – |
| TLR8 + | 0 | 29 | TLR8 + | 1.2 | 27.8 |
| TLR8 – | 6 | 107 | TLR8 – | 4.8 | 108.2 |
| **Pooled data** | TLR7 + | TLR7 – | | TLR7 + | TLR7 – |
| TLR8 + | 4 | 99 | TLR8 + | 6.5 | 96.5 |
| TLR8 – | 54 | 766 | TLR8 – | 51.5 | 768.5 |

#### B. Men with Klinefelter syndrome

**Observed frequencies** / **Expected frequencies**

| **K2** | TLR7 + | TLR7 – | | TLR7 + | TLR7 – |
|---|---|---|---|---|---|
| TLR8 + | 0 | 29 | TLR8 + | 1.2 | 27.8 |
| TLR8 – | 6 | 113 | TLR8 – | 4.8 | 114.2 |

| **K3** | TLR7 + | TLR7 – | | TLR7 + | TLR7 – |
|---|---|---|---|---|---|
| TLR8 + | 1 | 27 | TLR8 + | 2.4 | 25.6 |
| TLR8 – | 7 | 58 | TLR8 – | 5.6 | 59.4 |

| **K4** | TLR7 + | TLR7 – | | TLR7 + | TLR7 – |
|---|---|---|---|---|---|
| TLR8 + | 1 | 36 | TLR8 + | 1.4 | 35.6 |
| TLR8 – | 5 | 112 | TLR8 – | 4.6 | 112.4 |

| **K5** | TLR7 + | TLR7 – | | TLR7 + | TLR7 – |
|---|---|---|---|---|---|
| TLR8 + | 0 | 9 | TLR8 + | 0.7 | 8.3 |
| TLR8 – | 11 | 129 | TLR8 – | 10.3 | 129.7 |

| **K11** | TLR7 + | TLR7 – | | TLR7 + | TLR7 – |
|---|---|---|---|---|---|
| TLR8 + | 1 | 8 | TLR8 + | 0.6 | 8.4 |
| TLR8 – | 9 | 134 | TLR8 – | 9.4 | 133.6 |

| **Pooled data** | TLR7 + | TLR7 – | | TLR7 + | TLR7 – |
|---|---|---|---|---|---|
| TLR8 + | 3 | 109 | TLR8 + | 6.6 | 105.4 |
| TLR8 – | 38 | 546 | TLR8 – | 34.4 | 549.6 |

**S3 Data. RNA FISH analysis of CD14⁺ monocytes, total cells.** Left, observed frequencies: cell counts cross-classified as 2×2 contingency tables depending on the presence of transcriptional foci for *TLR7* or *TLR8*, regardless of Xa markers (Any X plots). Right, the corresponding expected frequencies, computed under the null hypothesis of mutually independent transcription of the two genes. Donor IDs in red.

#### A. Normal men

| Observed frequencies | | | Expected frequencies | | |
|---|---|---|---|---|---|
| **M366** | TLR7 + | TLR7 – | | TLR7 + | TLR7 – |
| TLR8 + | 5 | 77 | TLR8 + | 17.9 | 64.1 |
| TLR8 – | 50 | 120 | TLR8 – | 37.1 | 132.9 |
| **M387** | TLR7 + | TLR7 – | | TLR7 + | TLR7 – |
| TLR8 + | 6 | 63 | TLR8 + | 15.8 | 53.2 |
| TLR8 – | 60 | 160 | TLR8 – | 50.2 | 169.8 |
| **M235** | TLR7 + | TLR7 – | | TLR7 + | TLR7 – |
| TLR8 + | 4 | 73 | TLR8 + | 9.5 | 67.5 |
| TLR8 – | 22 | 112 | TLR8 – | 16.5 | 117.5 |
| **M525** | TLR7 + | TLR7 – | | TLR7 + | TLR7 – |
| TLR8 + | 7 | 35 | TLR8 + | 10.5 | 31.5 |
| TLR8 – | 68 | 190 | TLR8 – | 64.5 | 193.5 |
| **M657** | TLR7 + | TLR7 – | | TLR7 + | TLR7 – |
| TLR8 + | 7 | 50 | TLR8 + | 9.7 | 47.3 |
| TLR8 – | 44 | 198 | TLR8 – | 41.3 | 200.7 |
| **M660** | TLR7 + | TLR7 – | | TLR7 + | TLR7 – |
| TLR8 + | 5 | 65 | TLR8 + | 11.5 | 58.5 |
| TLR8 – | 34 | 133 | TLR8 – | 27.5 | 139.5 |
| **M661** | TLR7 + | TLR7 – | | TLR7 + | TLR7 – |
| TLR8 + | 7 | 66 | TLR8 + | 6.9 | 66.1 |
| TLR8 – | 22 | 210 | TLR8 – | 22.1 | 209.9 |

#### B. Women

**Observed frequencies** / **Expected frequencies**

| **F489** | TLR7 + | TLR7 – | | TLR7 + | TLR7 – |
|---|---|---|---|---|---|
| TLR8 + | 49 | 26 | TLR8 + | 22.1 | 52.9 |
| TLR8 – | 79 | 280 | TLR8 – | 105.9 | 253.1 |

| **F496** | TLR7 + | TLR7 – | | TLR7 + | TLR7 – |
|---|---|---|---|---|---|
| TLR8 + | 52 | 46 | TLR8 + | 32.6 | 65.4 |
| TLR8 – | 44 | 147 | TLR8 – | 63.4 | 127.6 |

| **F620** | TLR7 + | TLR7 – | | TLR7 + | TLR7 – |
|---|---|---|---|---|---|
| TLR8 + | 34 | 69 | TLR8 + | 21.2 | 81.8 |
| TLR8 – | 21 | 143 | TLR8 – | 33.8 | 130.2 |

| **F438** | TLR7 + | TLR7 – | | TLR7 + | TLR7 – |
|---|---|---|---|---|---|
| TLR8 + | 43 | 64 | TLR8 + | 31.9 | 75.1 |
| TLR8 – | 35 | 120 | TLR8 – | 46.1 | 108.9 |

| **F258** | TLR7 + | TLR7 – | | TLR7 + | TLR7 – |
|---|---|---|---|---|---|
| TLR8 + | 27 | 29 | TLR8 + | 21.9 | 34.1 |
| TLR8 – | 59 | 105 | TLR8 – | 64.1 | 99.9 |

| **F431** | TLR7 + | TLR7 – | | TLR7 + | TLR7 – |
|---|---|---|---|---|---|
| TLR8 + | 33 | 26 | TLR8 + | 21.3 | 37.7 |
| TLR8 – | 34 | 93 | TLR8 – | 45.7 | 81.3 |

#### C. Men with Klinefelter syndrome

**Observed frequencies** / **Expected frequencies**

| **K2** | TLR7 + | TLR7 – | | TLR7 + | TLR7 – |
|---|---|---|---|---|---|
| TLR8 + | 35 | 60 | TLR8 + | 19.0 | 76.0 |
| TLR8 – | 18 | 152 | TLR8 – | 34.0 | 136.0 |

| **K3** | TLR7 + | TLR7 – | | TLR7 + | TLR7 – |
|---|---|---|---|---|---|
| TLR8 + | 27 | 79 | TLR8 + | 20.7 | 85.3 |
| TLR8 – | 30 | 156 | TLR8 – | 36.3 | 149.7 |

| **K4** | TLR7 + | TLR7 – | | TLR7 + | TLR7 – |
|---|---|---|---|---|---|
| TLR8 + | 22 | 83 | TLR8 + | 15.4 | 89.6 |
| TLR8 – | 27 | 202 | TLR8 – | 33.6 | 195.4 |

| **K11** | TLR7 + | TLR7 – | | TLR7 + | TLR7 – |
|---|---|---|---|---|---|
| TLR8 + | 36 | 54 | TLR8 + | 15.7 | 74.3 |
| TLR8 – | 26 | 239 | TLR8 – | 46.3 | 218.7 |

| **K5** | TLR7 + | TLR7 – | | TLR7 + | TLR7 – |
|---|---|---|---|---|---|
| TLR8 + | 31 | 57 | TLR8 + | 17.9 | 70.1 |
| TLR8 – | 30 | 182 | TLR8 – | 43.1 | 168.9 |

**S4 Data. RNA FISH analysis of CD14⁺ monocytes, Xa-positive cells.** Left, observed frequencies: cell counts cross-classified as 2×2 contingency tables depending on the presence of transcriptional foci for *TLR7* or *TLR8*. Only those cells positive for the Xa marker probe (Xa⁺) were counted. Right, the corresponding expected frequencies, computed under the assumption of mutually independent transcription of the two genes. Donor IDs in red.

#### A. Normal men

| Observed frequencies | | | Expected frequencies | | |
|---|---|---|---|---|---|
| **M848** | TLR7 + | TLR7 – | | TLR7 + | TLR7 – |
| TLR8 + | 6 | 40 | TLR8 + | 12.1 | 33.9 |
| TLR8 – | 21 | 36 | TLR8 – | 14.9 | 42.1 |
| **M387** | TLR7 + | TLR7 – | | TLR7 + | TLR7 – |
| TLR8 + | 4 | 44 | TLR8 + | 16.7 | 31.3 |
| TLR8 – | 30 | 20 | TLR8 – | 17.3 | 32.7 |
| **M331** | TLR7 + | TLR7 – | | TLR7 + | TLR7 – |
| TLR8 + | 6 | 36 | TLR8 + | 16.1 | 25.9 |
| TLR8 – | 37 | 33 | TLR8 – | 26.9 | 43.1 |
| **M235** | TLR7 + | TLR7 – | | TLR7 + | TLR7 – |
| TLR8 + | 5 | 54 | TLR8 + | 13.6 | 45.4 |
| TLR8 – | 18 | 23 | TLR8 – | 9.4 | 31.6 |
| **M660** | TLR7 + | TLR7 – | | TLR7 + | TLR7 – |
| TLR8 + | 5 | 46 | TLR8 + | 22.4 | 28.6 |
| TLR8 – | 52 | 27 | TLR8 – | 34.6 | 44.4 |

#### B. Women

| Observed frequencies | | | Expected frequencies | | |
|---|---|---|---|---|---|
| **F489** | TLR7 + | TLR7 – | | TLR7 + | TLR7 – |
| TLR8 + | 34 | 15 | TLR8 + | 27.8 | 21.2 |
| TLR8 – | 21 | 27 | TLR8 – | 27.2 | 20.8 |
| **F620** | TLR7 + | TLR7 – | | TLR7 + | TLR7 – |
| TLR8 + | 20 | 22 | TLR8 + | 12.6 | 29.4 |
| TLR8 – | 10 | 48 | TLR8 – | 17.4 | 40.6 |
| **F438** | TLR7 + | TLR7 – | | TLR7 + | TLR7 – |
| TLR8 + | 34 | 46 | TLR8 + | 29.8 | 50.2 |
| TLR8 – | 11 | 30 | TLR8 – | 15.2 | 25.8 |
| **F258** | TLR7 + | TLR7 – | | TLR7 + | TLR7 – |
| TLR8 + | 23 | 5 | TLR8 + | 19.5 | 8.5 |
| TLR8 – | 14 | 11 | TLR8 – | 17.5 | 7.5 |
| **F431** | TLR7 + | TLR7 – | | TLR7 + | TLR7 – |
| TLR8 + | 32 | 16 | TLR8 + | 27.3 | 20.7 |
| TLR8 – | 14 | 19 | TLR8 – | 18.7 | 14.3 |

#### C. Men with Klinefelter syndrome

**Observed frequencies**

| **K2** | TLR7 + | TLR7 – |
|---|---|---|
| TLR8 + | 33 | 26 |
| TLR8 – | 7 | 21 |

| **K3** | TLR7 + | TLR7 – |
|---|---|---|
| TLR8 + | 18 | 36 |
| TLR8 – | 21 | 15 |

| **K4** | TLR7 + | TLR7 – |
|---|---|---|
| TLR8 + | 16 | 38 |
| TLR8 – | 12 | 26 |

| **K11** | TLR7 + | TLR7 – |
|---|---|---|
| TLR8 + | 27 | 29 |
| TLR8 – | 13 | 21 |

| **K5** | TLR7 + | TLR7 – |
|---|---|---|
| TLR8 + | 18 | 41 |
| TLR8 – | 24 | 17 |

**Expected frequencies**

| **K2** | TLR7 + | TLR7 – |
|---|---|---|
| TLR8 + | 27.1 | 31.9 |
| TLR8 – | 12.9 | 15.1 |

| **K3** | TLR7 + | TLR7 – |
|---|---|---|
| TLR8 + | 23.4 | 30.6 |
| TLR8 – | 15.6 | 20.4 |

| **K4** | TLR7 + | TLR7 – |
|---|---|---|
| TLR8 + | 16.4 | 37.6 |
| TLR8 – | 11.6 | 26.4 |

| **K11** | TLR7 + | TLR7 – |
|---|---|---|
| TLR8 + | 24.9 | 31.1 |
| TLR8 – | 15.1 | 18.9 |

| **K5** | TLR7 + | TLR7 – |
|---|---|---|
| TLR8 + | 24.8 | 34.2 |
| TLR8 – | 17.2 | 23.8 |

**S5 Data. Xa versus Xi comparison for patterns of *TLR7* and *TLR8* transcription.** The 3×2 tables show *TLR7*$^+$ *TLR8*$^+$, *TLR7*$^-$ *TLR8*$^+$, and *TLR7*$^+$ *TLR8*$^-$ cell counts in the RNA FISH data for the Xa and the Xi of monocytes from female and Klinefelter syndrome male donors; p-values from Monte Carlo $\chi^2$ tests with $10^6$ replications. Donor IDs in red.

#### A. Women.

| | | Xa | Xi | p-value |
|---|---|---|---|---|
| **F620** | | | | |
| | TLR7+ TLR8+ | 20 | 0 | 0.00060 |
| | TLR7– TLR8+ | 22 | 19 | |
| | TLR7+ TLR8– | 10 | 3 | |
| **F489** | | | | |
| | TLR7+ TLR8+ | 34 | 1 | 0.0013 |
| | TLR7– TLR8+ | 15 | 10 | |
| | TLR7+ TLR8– | 21 | 6 | |
| **F438** | | | | |
| | TLR7+ TLR8+ | 34 | 1 | 0.000048 |
| | TLR7– TLR8+ | 46 | 26 | |
| | TLR7+ TLR8– | 11 | 12 | |
| **F431** | | | | |
| | TLR7+ TLR8+ | 32 | 1 | 0.000074 |
| | TLR7– TLR8+ | 16 | 5 | |
| | TLR7+ TLR8– | 14 | 14 | |
| **F258** | | | | |
| | TLR7+ TLR8+ | 23 | 0 | 0.000001 |
| | TLR7– TLR8+ | 5 | 29 | |
| | TLR7+ TLR8– | 14 | 6 | |

#### B. Men with Klinefelter syndrome.

| | | Xa | Xi | p-value |
|---|---|---|---|---|
| **K2** | | | | |
| | TLR7+ TLR8+ | 33 | 0 | 0.000001 |
| | TLR7– TLR8+ | 26 | 29 | |
| | TLR7+ TLR8– | 7 | 6 | |
| **K3** | | | | |
| | TLR7+ TLR8+ | 18 | 1 | 0.0065 |
| | TLR7– TLR8+ | 36 | 27 | |
| | TLR7+ TLR8– | 21 | 7 | |
| **K4** | | | | |
| | TLR7+ TLR8+ | 16 | 1 | 0.0030 |
| | TLR7– TLR8+ | 38 | 36 | |
| | TLR7+ TLR8– | 12 | 5 | |
| **K5** | | | | |
| | TLR7+ TLR8+ | 18 | 0 | 0.021 |
| | TLR7– TLR8+ | 41 | 9 | |
| | TLR7+ TLR8– | 24 | 11 | |
| **K11** | | | | |
| | TLR7+ TLR8+ | 27 | 1 | 0.0051 |
| | TLR7– TLR8+ | 29 | 8 | |
| | TLR7+ TLR8– | 13 | 9 | |

**S6 Data. RNA FISH analysis of CD4⁺ T lymphocytes from women and normal men.** Left, observed frequencies: cell counts cross-classified as 2×2 contingency tables depending on the presence of transcriptional foci for *TLR7* or *TLR8*. Right, expected frequencies, computed from the observed frequencies under the assumption of mutually independent transcription of the two genes. Donor IDs in red. The any X data (**A**, **C**) corresponds to cells scored regardless of the hybridization with the Xa marker probe. In the $Xa^{+}$ data (**B**, **D**), only cells positive for the Xa marker probe were scored.

#### A. Women, any X

| Observed frequencies | | | Expected frequencies | | |
|---|---|---|---|---|---|
| **F438** | TLR7 + | TLR7 – | | TLR7 + | TLR7 – |
| TLR8 + | 47 | 38 | TLR8 + | 19.1 | 65.9 |
| TLR8 – | 24 | 207 | TLR8 – | 51.9 | 179.1 |
| **F557** | TLR7 + | TLR7 – | | TLR7 + | TLR7 – |
| TLR8 + | 54 | 53 | TLR8 + | 26.0 | 81.0 |
| TLR8 – | 20 | 178 | TLR8 – | 48.0 | 150.0 |
| **F595** | TLR7 + | TLR7 – | | TLR7 + | TLR7 – |
| TLR8 + | 44 | 65 | TLR8 + | 20.8 | 88.2 |
| TLR8 – | 18 | 198 | TLR8 – | 41.2 | 174.8 |
| **F619** | TLR7 + | TLR7 – | | TLR7 + | TLR7 – |
| TLR8 + | 19 | 66 | TLR8 + | 11.8 | 73.2 |
| TLR8 – | 20 | 175 | TLR8 – | 27.2 | 167.8 |
| **F689** | TLR7 + | TLR7 – | | TLR7 + | TLR7 – |
| TLR8 + | 45 | 45 | TLR8 + | 15.5 | 74.5 |
| TLR8 – | 8 | 210 | TLR8 – | 37.5 | 180.5 |

#### B. Women, $Xa^{+}$

| Observed frequencies | | | Expected frequencies | | |
|---|---|---|---|---|---|
| **F438** | TLR7 + | TLR7 – | | TLR7 + | TLR7 – |
| TLR8 + | 45 | 9 | TLR8 + | 34.6 | 19.4 |
| TLR8 – | 19 | 27 | TLR8 – | 29.4 | 16.6 |
| **F557** | TLR7 + | TLR7 – | | TLR7 + | TLR7 – |
| TLR8 + | 41 | 19 | TLR8 + | 28.2 | 31.8 |
| TLR8 – | 6 | 34 | TLR8 – | 18.8 | 21.2 |
| **F595** | TLR7 + | TLR7 – | | TLR7 + | TLR7 – |
| TLR8 + | 35 | 33 | TLR8 + | 32.3 | 35.7 |
| TLR8 – | 13 | 20 | TLR8 – | 15.7 | 17.3 |
| **F619** | TLR7 + | TLR7 – | | TLR7 + | TLR7 – |
| TLR8 + | 18 | 38 | TLR8 + | 19.6 | 36.4 |
| TLR8 – | 17 | 27 | TLR8 – | 15.4 | 28.6 |
| **F689** | TLR7 + | TLR7 – | | TLR7 + | TLR7 – |
| TLR8 + | 44 | 40 | TLR8 + | 40.8 | 43.2 |
| TLR8 – | 8 | 15 | TLR8 – | 11.2 | 11.8 |

#### C. Normal men, any X

**Observed frequencies** / **Expected frequencies**

| **M347** | TLR7 + | TLR7 – | | TLR7 + | TLR7 – |
|---|---|---|---|---|---|
| TLR8 + | 15 | 56 | TLR8 + | 17.1 | 53.9 |
| TLR8 – | 66 | 200 | TLR8 – | 63.9 | 202.1 |

| **M605** | TLR7 + | TLR7 – | | TLR7 + | TLR7 – |
|---|---|---|---|---|---|
| TLR8 + | 6 | 81 | TLR8 + | 10.5 | 76.5 |
| TLR8 – | 31 | 190 | TLR8 – | 26.5 | 194.5 |

| **M712** | TLR7 + | TLR7 – | | TLR7 + | TLR7 – |
|---|---|---|---|---|---|
| TLR8 + | 8 | 76 | TLR8 + | 9.2 | 74.8 |
| TLR8 – | 31 | 240 | TLR8 – | 29.8 | 241.2 |

| **M720** | TLR7 + | TLR7 – | | TLR7 + | TLR7 – |
|---|---|---|---|---|---|
| TLR8 + | 8 | 56 | TLR8 + | 7.6 | 56.4 |
| TLR8 – | 36 | 270 | TLR8 – | 36.4 | 269.6 |

| **M700** | TLR7 + | TLR7 – | | TLR7 + | TLR7 – |
|---|---|---|---|---|---|
| TLR8 + | 4 | 31 | TLR8 + | 7.1 | 27.9 |
| TLR8 – | 59 | 218 | TLR8 – | 55.9 | 221.1 |

#### D. Normal men, Xa$^+$

**Observed frequencies** / **Expected frequencies**

| **M347** | TLR7 + | TLR7 – | | TLR7 + | TLR7 – |
|---|---|---|---|---|---|
| TLR8 + | 11 | 16 | TLR8 + | 9 | 18 |
| TLR8 – | 23 | 52 | TLR8 – | 25 | 50 |

| **M605** | TLR7 + | TLR7 – | | TLR7 + | TLR7 – |
|---|---|---|---|---|---|
| TLR8 + | 6 | 47 | TLR8 + | 10.1 | 42.9 |
| TLR8 – | 13 | 34 | TLR8 – | 8.9 | 38.1 |

| **M712** | TLR7 + | TLR7 – | | TLR7 + | TLR7 – |
|---|---|---|---|---|---|
| TLR8 + | 6 | 36 | TLR8 + | 11.2 | 30.8 |
| TLR8 – | 21 | 38 | TLR8 – | 15.8 | 43.2 |

| **M720** | TLR7 + | TLR7 – | | TLR7 + | TLR7 – |
|---|---|---|---|---|---|
| TLR8 + | 6 | 36 | TLR8 + | 10.9 | 31.1 |
| TLR8 – | 20 | 38 | TLR8 – | 15.1 | 42.9 |

| **M700** | TLR7 + | TLR7 – | | TLR7 + | TLR7 – |
|---|---|---|---|---|---|
| TLR8 + | 4 | 31 | TLR8 + | 16.8 | 18.2 |
| TLR8 – | 44 | 21 | TLR8 – | 31.2 | 33.8 |
